# Timing of antiretroviral treatment initiation and seropositivity to measles virus among children living with HIV in rural Zambia

**DOI:** 10.64898/2026.03.09.26347987

**Authors:** Catherine G. Sutcliffe, Saki Takahashi, Amanda A. Finney, Mutinta Hamahuwa, Nkumbula Moyo, Amy K. Winter, Hellen K. Matakala, Mathias Muleka, Passwell Munachoonga, Francis Hamangaba, Philip E. Thuma, William J. Moss

## Abstract

**Background:** Combination antiretroviral therapy (cART) reduces morbidity among children living with HIV (CHIV) but does not restore measles immunity. Many CHIV now initiate cART before measles vaccination but the impact on measles-specific immunity is not understood. This study compared measles antibodies between CHIV initiating cART before and after 9 months of age in Zambia.

**Methods:** This retrospective study was nested within an open cohort study of CHIV and community-based malaria studies in Macha, Zambia conducted from 2007-2020. Samples were tested for measles IgG antibodies from CHIV defined by age at cART initiation (aged 0-8, 9-23, and 24-59 months) and age-matched community-based controls. A cross-sectional analysis compared antibody concentrations between CHIV groups after 6-12 months of cART and aged >12 months and controls. A longitudinal analysis evaluated antibody trajectories from 9 months of age through 48 months of cART and during the 2010-11 measles outbreak.

**Results:** In the cross-sectional analysis, 31.9% of 301 CHIV were seropositive after 6-12 of cART, with no significant difference by group (0-8: 30.8%; 9-23: 37.0%; 24-59 months: 26.6%). Within each CHIV group, the proportion seropositive was significantly lower than age-matched controls. Among 242 CHIV included in the longitudinal analysis, 46-56% had an antibody response, with an estimated time to seroreversion of 1.8-3.2 years. Among CHIV followed in 2010-11, 50% seroconverted or boosted; the estimated time to seroreversion was 2.1 years.

**Conclusions:** Low levels of measles antibodies were found among CHIV, even when cART was started before 9 months of age, supporting a recommendation that CHIV, including infants, would benefit from revaccination.

## Introduction

Untreated children living with HIV (CHIV) generally have a poor response to primary vaccination and more rapid waning immunity after vaccination,^1^ potentially leaving them susceptible to vaccine-preventable diseases, including measles. Combination antiretroviral therapy (cART) is effective in reducing HIV-related morbidity and mortality through suppression of viral replication and restoration of immune function.^2^ However, immune reconstitution in children is primarily through generation of naïve T-cells,^3^ therefore vaccine-induced immunity is not restored after cART initiation.^4^ If revaccinated with measles-containing vaccine (MCV), CHIV receiving cART are able to mount a good antibody response, although waning immunity may still occur^4^ due to persistent B-cell abnormalities.^5^ Based on this evidence, the World Health Organization (WHO) recommends that CHIV receive an additional dose of MCV either when immune reconstitution is achieved or 6-12 months after cART initiation.^6, 7^

Over the past two decades, the increasing recognition that early cART initiation reduces morbidity^8^ resulted in recommendations for a test-and-treat strategy for infants in 2008 and all CHIV by 2016.^9, 10^ Following increased access to early infant diagnosis, the age at cART initiation decreased in the African region.^11^ With cART initiation in infancy, CD4 T-cell counts are stabilized^12^ and total memory B-cell percentages are preserved.^13^ However, the impact of cART initiation in infancy on antigen-specific immunity is not fully understood as few studies have been conducted.^13, 14^ This is particularly important for MCV given its administration at 9 months of age when many CHIV may have been diagnosed and initiated cART before receiving the first dose of MCV (MCV1). With introduction of a second routine dose of MCV (MCV2) at 15-18 months of age, CHIV initiating cART in infancy also may receive a second dose after immune reconstitution. Understanding the impact of this change in the relative timing of measles vaccination and cART on immunity to measles is important for providing CHIV with optimal care and potentially mitigating the impact of measles-susceptible CHIV on regional measles elimination efforts.^15^ When WHO recommended revaccination, there was insufficient evidence to include CHIV initiating cART before MCV1.^6, 7^

This study was conducted to evaluate the short and long-term impact of cART initiation before 9 months of age on levels of measles antibodies among CHIV younger than 5 years of age in Zambia.

## Materials and Methods

### Overview

This retrospective study was nested within two studies conducted in the catchment area of Macha Hospital in Choma District, Southern Province, Zambia: 1) the Pediatric Antiretroviral Treatment (PART) study, a clinical cohort of CHIV; and 2) a set of community-based studies conducted through the International Center of Excellence for Malaria Research (ICEMR) (SDC 1). Stored samples from CHIV (PART study) and control children (ICEMR studies) were tested for measles IgG antibodies for inclusion in cross-sectional and longitudinal analyses (SDC 2). The cross-sectional analysis included samples from CHIV 6-12 months after cART initiation and age-matched control children to evaluate the short-term impact of cART initiation before 9 months of age. The longitudinal analysis included samples from CHIV up to four years after cART initiation and modelled antibody decay kinetics to evaluate the long-term impact of cART initiation before 9 months of age.

### Ethics statement

These studies were approved by the Institutional Review Boards at Johns Hopkins Bloomberg School of Public Health (#447and #3467), the University of Zambia (002-05-07), and the Tropical Diseases Research Center (TDRC/ERC/2010/14/11), and the National Health Research Authority in Zambia. For all studies, informed consent included permission to use the samples for approved research.

### Setting

The Macha area is populated primarily by subsistence farmers living in small, scattered homesteads. The HIV prevalence in Southern Province and Choma District was 12.5% and 17.2%, respectively, in 2016.^16, 17^ Immunization coverage was high in Southern Province and improved over time: the proportion of children aged 12-36 months receiving MCV1 (recommended at 9 months of age) increased from 86.0% in 2013 to 94.5% in 2018;^18, 19^ in Choma District, 95.7% of children 9-60 months received MCV1 in 2020.^20^ A second dose of MCV (MCV2) was introduced at 18 months of age in 2013 and coverage was 57.6% among children aged 24-36 months in 2018.^18^ To increase vaccination coverage in the face of measles outbreaks, supplemental immunization activities (SIA) were conducted in 2007, 2010, 2012 and 2016 (SDC 1).

### PART study – source of CHIV samples

An open HIV clinical cohort study was conducted at the Macha HIV clinic from 2007-2023.^21^ CHIV 0-15 years of age were eligible for enrollment. Written informed consent was obtained from the primary caregiver and assent from children 12-15 years of age. Children had study visits every three months that included a questionnaire, height and weight measures, and biannual collection of a venous blood sample. The blood sample was processed and stored frozen as plasma (-80°C) and as a dried blood spot (DBS) card (-20°C). Clinical information was abstracted from the medical record. For children enrolled 2007-2012, the under-five card was reviewed at enrollment (if available) to document receipt of routine childhood vaccines, including MCV.

### ICEMR studies – source of controls samples

Annual cross-sectional surveys were conducted in the Macha area from 2008-2013 to determine changes in malaria parasite prevalence.^22^ Households were randomly selected bi-monthly and all household members were eligible. From 2015-2017, a malaria reactive test-and-treat study was conducted in the Macha area.^23^ When an individual tested positive for malaria at a health facility, residents within 250 m of the index case household were enrolled. In 2018, a longitudinal cohort study evaluating malaria transmission was initiated in a community in the catchment area. All households in the study area were invited to participate and all household members were eligible.

For each study, written informed consent was obtained from each adult or parent. At study visits, a questionnaire was administered to collect information on demographics, symptoms, and use of medications (including cART), and a blood sample was collected by finger prick on DBS cards which were stored at −20°C. Vaccination data were not available.

### Sample selection

<u>CHIV – cross-sectional analysis:</u> PART participants were eligible if they initiated cART <5 years of age, and 2) had a DBS or plasma sample available 6-12 months after cART initiation between 2007 and 2020. The first available sample >12 months of age (to allow time to receive MCV1) was selected. <u>CHIV – longitudinal analysis:</u> PART participants were eligible if 1) they initiated cART <5 years of age, and had at least two DBS or plasma samples available after cART initiation. The last available pre-cART sample and all post-cART samples up to 48 months were selected. <u>Controls – cross-sectional analysis:</u> Samples from participants aged 0-5 years in the ICEMR cross-sectional surveys, reactive test–and-treat study, and baseline visit of the longitudinal study were eligible if they did not self-report receiving cART. Samples were selected in one year age bands to match CHIV age at sample collection (ratio of 3:1).

### Laboratory testing

DBS and plasma samples were tested for anti-measles virus IgG antibodies in the Clinical Research Laboratory at Macha Research Trust using the Euroimmun Anti-Measles Virus ELISA (Euroimmun AG, Lübeck, Germany) according to manufacturer recommendations.^24^ Because of concerns about assay sensitivity,^25^ results were adjusted using a correction factor derived from a comparison study between the Euroimmun ELISA and a Multiplex Bead Assay (MBA; SDC 3). After adjustment, samples with antibody concentration ≥153 mIU/mL were considered positive.^26^

Due to concerns about storage duration (range 1-13 years) and sample types (plasma and DBS), a comparison study between paired DBS and plasma samples from the PART study was conducted (SDC 4). No impact of storage duration was observed, but antibody concentrations from plasma were systematically higher than DBS and a correction factor was derived. Results from plasma samples were adjusted using the correction factor before applying the MBA correction factor.

### Statistical analysis

#### Definitions

CHIV were grouped according to timing of cART initiation in relation to presumed measles vaccination: 1) 0-8 months of age, presumed before MCV1 (Early cART[0-8m]); 2) 9-23 months of age, presumed shortly after MCV1 (Late cART[9-23m]); and 3) 24-59 months of age, presumed after MCV1 (Late cART[24-59m]).

#### Cross-sectional analysis

Descriptive statistics were used to summarize and compare the characteristics of CHIV overall and in each group and to compare measles antibody concentrations and proportion seropositive between CHIV groups and age-matched controls. As groups differed by age and sex, bivariable and multivariable prevalence ratios and 95% confidence intervals (CI) were estimated using Poisson regression with robust variance estimation. Among CHIV, correlates of seropositivity were also explored using Poisson regression with robust variance estimation. Characteristics marginally significant (p<0.2) in bivariable analysis were included in a multivariable model. To evaluate the impact of vaccination status on the results, a sensitivity analysis was conducted among CHIV with data on MCV status (SDC 5).

#### Longitudinal analysis: Post-MCV1 antibody decay

Children were included if they had a documented MCV1 date (n=66) or no vaccine card available (n=264; n=4 with vaccine card and no MCV date were excluded). To model antibody decay kinetics, time was calibrated to years since MCV1, as age at the first sample varied by cART group (SDC 6). The age of MCV1 receipt was recorded (SDC 7) or assumed to be 9 months. For individuals with documented MCV1, samples after receipt were included. Individuals were censored if there was evidence of additional MCV doses or infection, defined as either the time point before first observed seroconversion (seronegative to seropositive) or ≥2-fold increase in antibody concentration, to optimize sensitivity. The primary analysis was restricted to CHIV with ≥2 samples after applying censoring criteria (SDC 8).

We fit Bayesian growth (mixed effects) mixture models to the observed antibody trajectories and assumed that individuals were from a mixture of two latent serological groups: “non-responders” whose antibody concentrations at each time point were at the lower limit of detection (for any reason, including not being vaccinated, failing to respond to the vaccine, or waning), and “decayers” whose antibody concentrations decayed over time. We estimated the probability of being in either serological group for each cART group. Among the “decayers”, we incorporated a group-specific intercept and slope, and an individual-level random intercept. This model assumed a log-linear decay of antibody concentrations over time. For each group, we estimated the probability of being a “non-responder” and mean time to seroreversion after MCV among the “decayers” predicted to be seropositive at MCV receipt (SDC 9, Document). Sensitivity analyses were conducted using a ≥4-fold criteria for censoring (SDC 10) and including data from individuals with one observation (SDC 11).

#### 2010-2011 measles outbreak and vaccine campaign, and post-seroconversion decay

Zambia experienced a large measles outbreak between June 2010 and July 2011, with 37,582 cases reported including 1244 in Southern Province,^27^ and implemented a SIA campaign in July 2010 for children 9-47 months of age.^28^ Subsequent SIA campaigns occurred in September 2012, September 2016, and November 2020.^29^ To investigate evidence of these exposures to wild-type measles virus or vaccination, we examined antibody boosts, defined as seroconversion or a ≥4-fold increase in antibody concentration (a more stringent threshold to increase specificity). Individuals were allowed to experience multiple boosts during follow-up.

Additional analyses were conducted for the 2010-2011 outbreak. A 5-month buffer was added to define the outbreak period as January 2010 to December 2011. The analyses included individuals with ≥2 observations during the outbreak period, whose earliest time point was before August 2011 (i.e., end of outbreak) and latest time point after June 2010 (i.e., start of outbreak). The antibody trajectories of individuals experiencing seroconversion during the outbreak period were determined. Individuals were censored at either the time point before another observed seroconversion or ≥2-fold increase in the antibody concentration. A simple mixed effects model was fit to estimate the mean time to seroreversion (SDC 9).

## Results

### Cross-sectional analysis

Of the 472 CHIV initiating cART <5 years of age in the PART study, 301 were eligible for the analysis, including 39, 138, and 124 in the Early cART[0-8m], Late cART[9-23m], and Late cART[24-59m] group, respectively (Table 1). Measles antibody concentrations 6-12 months after cART and >12 months of age were low, with a median of 53 mIU/mL (interquartile range [IQR]: 16, 221). The proportion seropositive was only 31.9 % (Figure 1), including 30.8%, 37.0%, and 26.6% in the Early cART[0-8m], Late cART[9-23m], and Late cART[24-59m] group, respectively (p=0.20; Table 2). The results were not significantly different among CHIV with available MCV data or CHIV with documented receipt of MCV (SDC 5). Within each group, antibody concentrations and proportion seropositive were significantly lower than age-matched controls (63.5%, 75.7%, and 80.3% for the Early cART[0-8m], Late cART[9-23m], and Late cART[24-59m] controls, respectively) (Table 2; Figure 1).

**Figure 1.**
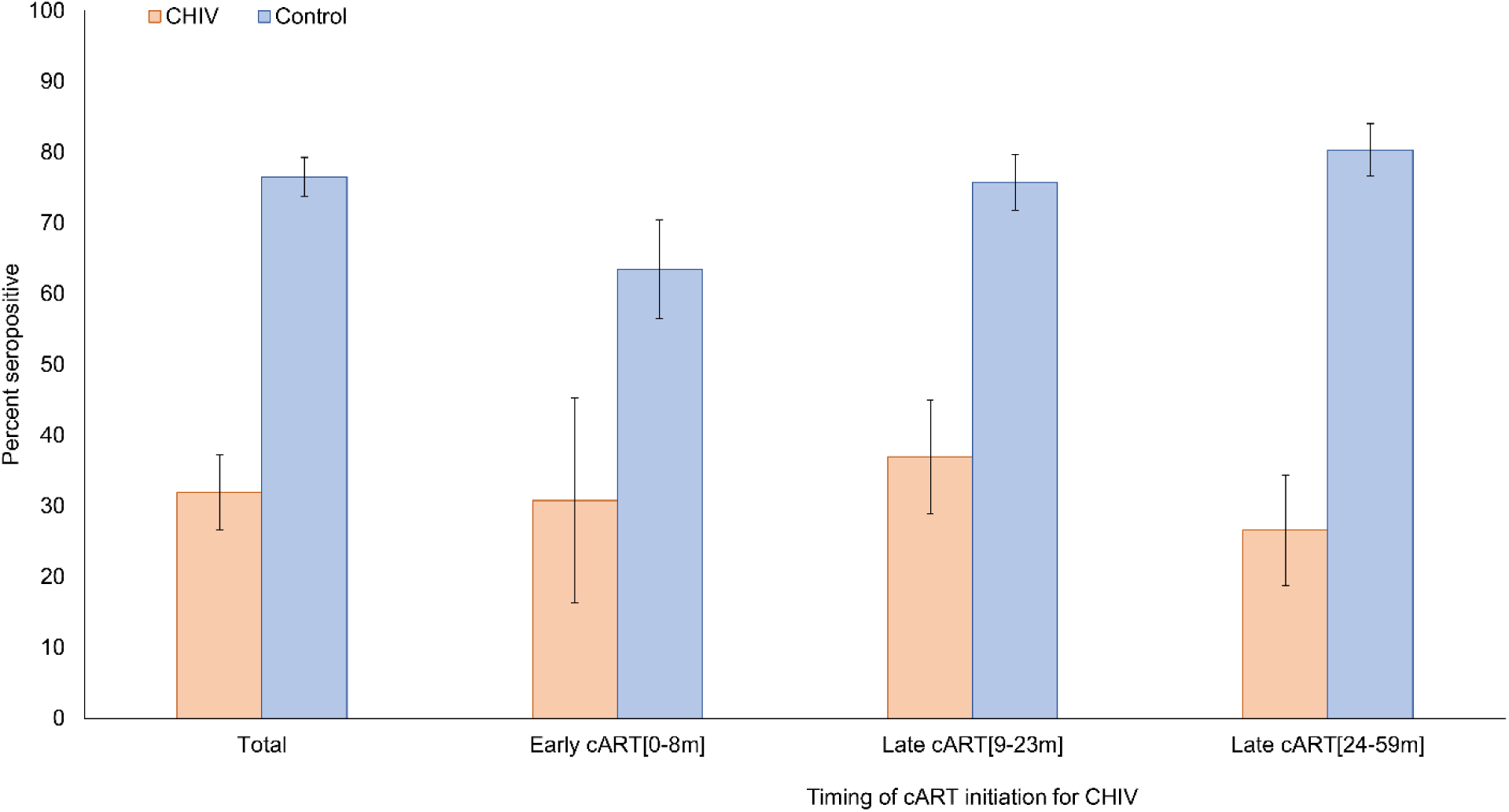
Measles seroprevalence among children living with HIV by timing of treatment initiation^a^ and age-matched controls. cART: combination antiretroviral therapy; CHIV: children living with HIV ^a^ Early cART[0-8m] defined as cART initiation 0-8 month of age; Late cART[9-23m] defined as cART initiation 9-23 months of age; Late cART[24-59m] defined as cART initiation 24-59 months of age

**Table 1.** Characteristics of children living with HIV overall and by timing of treatment initiation^a^.

|  | Total | Early cART<br>[0-8m] | Late cART<br>[9-23m] | Late cART<br>[24-59m] |  |
| --- | --- | --- | --- | --- | --- |
| **Characteristics at cART initiation** | **N=301** | **N=39** | **N=138** | **N=124** | **p-value<sup>h</sup>** |
| Age (years), median (IQR) | 1.7 (1.1,2.9) | 0.6 (0.5,0.7) | 1.4 (1.1,1.7) | 3.1 (2.5,4.2) | <0.001 |
| Female, N (%) | 147 (48.8) | 24 (61.5) | 73 (52.9) | 50 (40.3) | 0.03 |
| Single or double orphan<sup>b</sup> | 25 (8.3) | 1 (2.6) | 10 (7.2) | 14 (11.3) | 0.19 |
| SES percentile<sup>c</sup>, N (%) |  |  |  |  |  |
| 0-25% | 184 (61.1) | 24 (61.5) | 85 (61.6) | 75 (60.5) | 0.70 |
| 25-50% | 95 (31.6) | 12 (30.8) | 46 (33.3) | 37 (29.8) |  |
| 50-100% | 22 (7.3) | 3 (7.7) | 7 (5.1) | 12 (9.7) |  |
| CD4 T-cell percentage |  |  |  |  |  |
| Median (IQR) | 18.6 (13.0,24.6) | 23.5 (13.8,31.0) | 19.1 (15.2,25.2) | 17.1 (10.2,21.2) | <0.001 |
| Severe immunosuppression<sup>d</sup> | 145 (56.6) | 15 (55.6) | 69 (57.5) | 61 (56.0) | 0.98 |
| Missing | 45 |  |  |  |  |
| Height-for-age Z-score<sup>e</sup> |  |  |  |  |  |
| Median (IQR) | -2.9 (-3.9,-1.9) | -2.4 (-3.8,-1.6) | -3.2 (-4.0,-2.3) | -2.9 (-3.8,-1.9) | 0.27 |
| None/mild stunting | 59 (25.4) | 12 (42.9) | 22 (21.0) | 25 (25.3) | 0.18 |
| Moderate stunting | 62 (26.7) | 5 (17.9) | 28 (26.7) | 29 (29.3) |  |
| Severe stunting | 111 (47.8) | 11 (39.3) | 55 (52.4) | 45 (45.5) |  |
| Missing | 69 |  |  |  |  |
| Weight-for-age Z-score<sup>e</sup> |  |  |  |  |  |
| Median (IQR) | -1.8 (-2.9,-0.7) | -0.7 (-2.0,-0.1) | -2.1 (-3.1,-0.8) | -1.8 (-2.9,-1.1) | 0.002 |
| None/mild underweight | 154 (54.0) | 27 (75.0) | 64 (48.9) | 63 (53.4) | 0.04 |
| Moderate underweight | 65 (22.8) | 5 (13.9) | 29 (22.1) | 31 (26.3) |  |
| Severe underweight | 66 (23.2) | 4 (11.1) | 38 (29.0) | 24 (20.3) |  |
| Missing | 16 |  |  |  |  |
| Year of cART initiation, median (IQR) | 2011 (2010,2013) | 2011 (2010,2014) | 2011 (2010,2014) | 2011 (2009,2013) | 0.31 |
| cART combination regimen |  |  |  |  |  |
| ABC/3TC/NPV | 12 (4.0) | 1 (2.6) | 7 (5.2) | 4 (3.3) | <0.001 |
| AZT/3TC/NVP | 62 (20.9) | 12 (30.8) | 30 (22.2) | 20 (16.3) |  |
| D4T/3TC/NVP | 65 (21.9) | 12 (30.8) | 24 (17.8) | 29 (23.6) |  |
| ABC/3TC/EFV | 25 (8.4) | 0 (0.0) | 11 (8.1) | 14 (11.4) |  |
| AZT/3TC/EFV | 44 (14.8) | 2 (5.1) | 12 (8.9) | 30 (24.4) |  |
| D4T/3TC/EFV | 34 (11.4) | 2 (5.1) | 19 (14.1) | 13 (10.6) |  |
| (ABC or AZT)/3TC/LPV/r | 55 (18.5) | 10 (25.6) | 32 (23.7) | 13 (10.6) |  |
| Missing | 4 |  |  |  |  |
| **Characteristics at measles antibody testing** |  |  |  |  |  |
| Age (years), median (IQR) | 2.6 (2.0,3.7) | 1.4 (1.3,1.5) | 2.2 (1.9,2.6) | 4.0 (3.4,5.1) | <0.001 |
| Duration of cART (months), median (IQR) | 11.2 (8.9,11.9) | 10.7 (8.9,11.9) | 11.1 (8.9,11.9) | 11.4 (9.1,12.1) | 0.22 |
| CD4 T-cell percentage |  |  |  |  |  |
| Median (IQR) | 32.6 (26.1,38.7) | 30.9 (25.8,41.2) | 33.7 (26.2,38.9) | 32.0 (26.2,38.3) | 0.87 |
| Severe immunosuppression<sup>d</sup>, N (%) | 20 (7.4) | 3 (8.6) | 15 (12.2) | 2 (1.8) | 0.009 |
| Missing | 30 |  |  |  |  |
| Viral suppression<sup>f</sup>, N (%) | 177 (80.5) | 28 (75.7) | 82 (80.4) | 67 (82.7) | 0.67 |
| Missing | 81 |  |  |  |  |
| Height-for-age Z-score<sup>e</sup> |  |  |  |  |  |
| Median (IQR) | -2.7 (-3.7,-1.8) | -3.1 (-4.3,-1.9) | -2.9 (-3.9,-2.1) | -2.4 (-3.2,-1.6) | 0.005 |
| None/mild stunting | 81 (29.0) | 9 (25.0) | 28 (22.6) | 44 (37.0) | 0.03 |
| Moderate stunting | 82 (29.4) | 8 (22.2) | 36 (29.0) | 38 (31.9) |  |
| Severe stunting | 116 (41.6) | 19 (52.8) | 60 (48.4) | 37 (31.1) |  |
| Missing | 22 |  |  |  |  |
| Weight-for-age Z-score<sup>e</sup> |  |  |  |  |  |
| Median (IQR) | -1.2 (-2.0,-0.5) | -1.0 (-2.0,-0.3) | -1.2 (-1.9,-0.5) | -1.2 (-1.9,-0.6) | 0.84 |
| None/mild underweight | 217 (76.7) | 28 (75.7) | 97 (76.4) | 92 (77.3) | 0.74 |
| Moderate underweight | 38 (13.4) | 7 (18.9) | 17 (13.4) | 14 (11.8) |  |
| Severe underweight | 28 (9.9) | 2 (5.4) | 13 (10.2) | 13 (10.9) |  |
| Missing | 18 |  |  |  |  |
| Adherence >95%<sup>g</sup>, N (%) | 180 (73.8) | 22 (71.0) | 82 (73.9) | 76 (74.5) | 0.93 |
| Missing | 57 |  |  |  |  |

3TC: lamivudine; ABC: abacavir; AZT: zidovudine; cART: combination antiretroviral therapy; D4T: stavudine; EFV: efavirenz; LPV/r: lopinavir plus ritonavir; NVP: nevirapine; IQR: interquartile range; SD: standard deviation; SES: socioeconomic status
<sup>a</sup> Early cART[0-8m] defined as cART initiation 0-8 month of age; Late cART[9-23m] defined as cART initiation 9-23 months of age; Late cART[24-59m] defined as cART initiation 24-59 months of age
<sup>b</sup> Single/double orphan defined as either or both the child's mother or father deceased, as determined by caregiver report.
<sup>c</sup> Socioeconomic status defined based on the Demographic and Health Survey SES scale used in Zambia at the time the PART study started (*Zambia Central Statistical Office Lusaka. Zambia Demographic and Health Survey. Calverton, Maryland, USA: ORC and Macro International, Inc; 2003*). SES percentiles were based on the predetermined cutoffs (<25<sup>th</sup>=0-6; 26-50<sup>th</sup>=7-12; 51-75<sup>th</sup>=13-18; >75<sup>th</sup>=19-24).
<sup>d</sup> Based on WHO criteria for severe immunosuppression, using CD4 percentage and age at cART initiation (*WHO. Antiretroviral therapy of HIV infection in infants and children: towards universal access. Recommendations for a public health approach. Geneva, Switzerland: World Health Organization; 2006*).
<sup>e</sup> Based on WHO growth standards (https://www.who.int/toolkits/child-growth-standards). None/mild defined as a z-score $\geq -2$ ; moderate defined as a z-score $< -2$ and $\geq -3$ ; severe defined as a z-score $< -3$ .
<sup>f</sup> Viral suppression defined as a viral load <400 copies/mL.
<sup>g</sup> Adherence was measured by pill count and categorized as $\leq 95\%$ or $> 95\%$ based on the minimum value of all drugs counted.
<sup>h</sup> p-value based on Pearson's chi-square test for categorical variables and Kruskal-Wallis test for continuous variables.

**Table 2.**
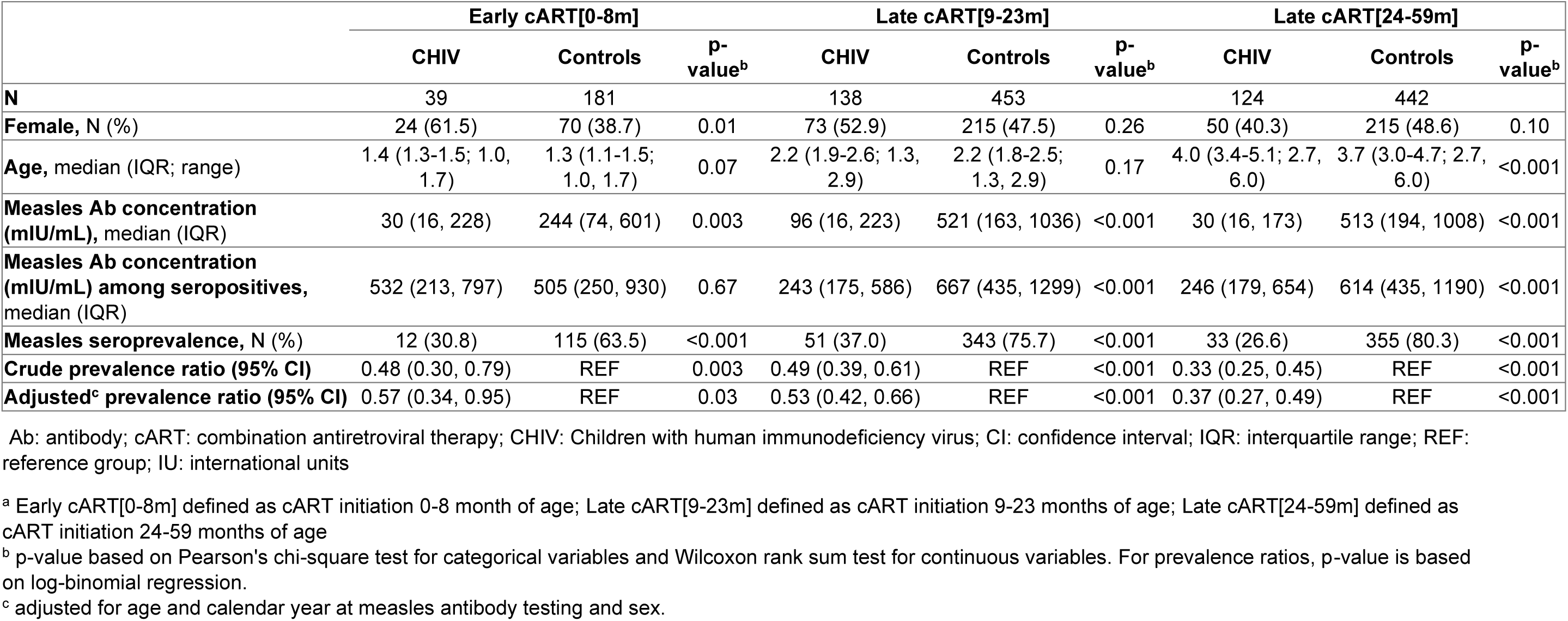
Characteristics and measles antibody concentrations of children living with HIV by timing of treatment initiation^a^ and age-matched controls.

Among CHIV, correlates of seropositivity were explored (SDC 12). Only single/double orphans and participants severely stunted at measles antibody testing were significantly more likely to be seropositive. In addition, seropositivity varied by calendar year and was significantly higher in 2011, 2013, 2017, and 2018-20 (SDC 12; SDC 13).

### Longitudinal analysis - Post-MCV1 antibody decay

Of the 242 CHIV (868 samples; Figure 2A) included, approximately half (44-54% by cART group) were “non-responders” (Table 3) and half were “decayers” (46-56%), most (67-84%) of whom were estimated to be seropositive at MCV receipt. Among decayers, there were no differences in the intercept (i.e., inferred antibody concentration at MCV1 receipt) by group. CHIV initiating cART the earliest (Early cART[0-8m]) had significantly faster antibody decay (-0.91 per year) compared to CHIV initiating cART at older ages (Late cART[9-23m]: -0.52 per year; Late cART[24-59m]: -0.46 per year) and thus a faster mean time to seroreversion (1.82 vs. 2.47 vs. 3.23 years, respectively). Importantly, CHIV across groups were predicted to serorevert 2-3 years after receiving MCV1.

**Figure 2.**
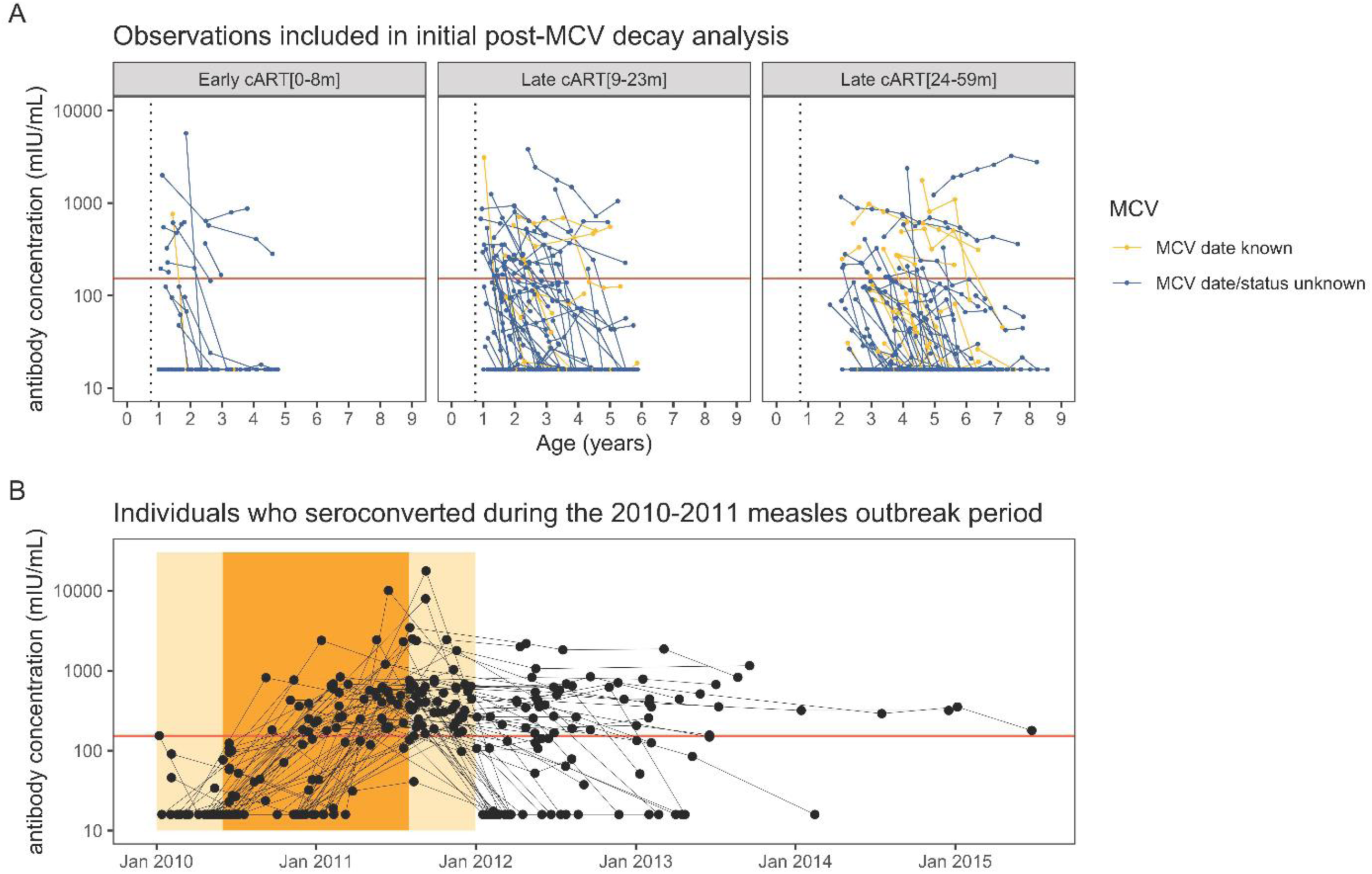
Longitudinal trajectories of measles antibody responses. **(A)** 242 individuals included in the initial post-MCV decay analysis. X-axis represents age in years, and y-axis represents adjusted antibody titer. Time points at and after censoring (defined as seroconversion or a ≥2-fold increase) are not shown. The columns indicate the ART group. Yellow circles indicate observations for individuals for whom the date of MCV receipt was recorded; blue circles indicate observations for individuals for whom date or confirmation of MCV receipt was unavailable and assumed to have occurred exactly at 9 months of age. The blue vertical dotted line on each panel indicates 9 months of age. The red horizontal line on each panel indicates the cutoff for seropositivity. **(B)** 59 individuals who seroconverted during the 2010-2011 measles outbreak period who had 1 or more observations after the outbreak period. X-axis represents calendar time, and y-axis represents adjusted antibody titer. Time points at and after censoring (defined as seroconversion or a ≥2-fold increase) are not shown. The orange shading represents the timing of the measles outbreak and SIA in June 2010-July 2011, and the light orange shading represents the 5-month buffer before and after this period. The red horizontal line indicates the cutoff for seropositivity. MCV: measles-containing vaccine

**Table 3:** Results of the post-MCV1 antibody decay analysis among children living with HIV.

| ART group (k)<sup>a</sup> | Probability of being a non-responder ($w_k$) | Probability of being a decayer ($1-w_k$) | ART group-specific intercept, among decayers ($\alpha_k$)<sup>b</sup> | ART group-specific slope, among decayers ($\beta_k$)<sup>c</sup> | Mean time to seroreversion, among decayers who are seropositive at the time of MCV1 receipt, years ($T_k$) | Proportion of decayers who are seropositive at the time of MCV1 receipt |
| --- | --- | --- | --- | --- | --- | --- |
| Early cART[0-8m] | 0.54 (0.37-0.71) | 0.46 (0.29-0.63) | 6.32 (5.53-7.04) | -0.91 (-1.18, -0.67) | 1.82 (1.39-2.45) | 0.84 (0.64-0.94) |
| Late cART[9-23m] | 0.44 (0.33-0.55) | 0.56 (0.45-0.67) | 5.60 (5.23-6.02) | -0.52 (-0.61, -0.43) | 2.47 (2.05-3.01) | 0.67 (0.55-0.78) |
| Late cART[24-59m] | 0.51 (0.41-0.62) | 0.48 (0.38-0.59) | 6.04 (5.52-6.54) | -0.46 (-0.55, -0.37) | 3.23 (2.70-3.88) | 0.78 (0.64-0.88) |

cART: combination antiretroviral therapy; MCV: measles-containing vaccine. The analysis includes children with at least 2 observations and censors children at seroconversion (seronegative to positive) or at $\geq 2$ -fold increase in antibody levels. Posterior median and 95% credible intervals are presented.
<sup>a</sup> Early cART[0-8m] defined as cART initiation 0-8 month of age; Late cART[9-23m] defined as cART initiation 9-23 months of age; Late cART[24-59m] defined as cART initiation 24-59 months of age
<sup>b</sup> The intercept represents the log antibody concentration at MCV1 receipt
<sup>c</sup> The slope represents the change in log antibody concentration per year

### Longitudinal analysis - 2010-2011 measles outbreak and vaccine campaign, and post-seroconversion decay

Among 195 CHIV with ≥1 antibody boost during follow-up (SDC 14), 260 boosting events were observed (SDC 15); 137, 51, and 7 CHIV had one, two, and three boosts, respectively. Assuming that boosts could occur at any time during follow-up, 43.0% (95% CI: 35.7%-50.7%) of eligible CHIV boosted in 2011, which was higher than in other years (SDC 15). Among 167 CHIV with ≥2 observations during the 2010-2011 outbreak period, 69 (41.3%) seroconverted (SDC 16). Among the 167 CHIV, no deaths were reported and 4 reported having ‘measles’ during the outbreak period, including 2 who seroconverted and 1 with an increase in antibody concentration that did not meet criteria for boosting. Among the 59 individuals who seroconverted and had ≥1 observation after the outbreak period (Figure 2B), the estimated mean time to seroreversion was comparable to the post-MCV1 decay analysis (posterior median: 2.1 years, 95% credible interval: 1.6-3.9 years).

## Discussion

Only one third of CHIV were measles seropositive within the first year of receiving cART with no significant differences by timing of cART initiation, even for those starting treatment before 9 months of age. Within each group, the proportion seropositive was significantly lower than age-matched controls. In longitudinal analyses, only one third of CHIV, regardless of timing of cART initiation, were estimated to be measles seropositive after receiving MCV1. Antibody levels declined rapidly, with seroreversion estimated within 2-3 years after measles vaccination.

These findings are consistent with prior studies from Zambia. Only 23% of CHIV aged 2-3 years in Lusaka in 2010 were measles seropositive before starting cART and no significant increase in measles antibody levels was observed up to 18 months later.^28^ In a nationally representative survey from Zambia in 2016, measles seroprevalence increased with age but was significantly lower among CHIV, most of whom were receiving cART.^30^ Among CHIV 0-9 years of age, only 47.2% were seropositive compared to 76.4% of HIV-uninfected children.^30^

Several studies of measles seropositivity found a benefit to initiating cART in the first year of life.^13, 14, 31–33^ In an Italian study of 70 CHIV with measles antibodies measured later in childhood (mean 6.8-15.8 years of age across groups), children initiating cART early in life (mean 6.8 months) had a higher measles seroprevalence (82%) than children initiating later in life (mean 7.4 years; 39%) and were more similar to uninfected controls (100%).^13^ In the Children with HIV Early Antiretroviral Therapy (CHER) trial in South Africa, CHIV in the deferred cART group, 71% of whom were receiving cART at MCV1 and all of whom had CD4 T-cell percentage ≥25% at cART initiation, had similar measles seroprevalence as controls at 2 (96.0% vs. 94.5%) and 4.5 (98.4% vs. 100%) years of age.^14, 33^ In this study, more than half of CHIV in the youngest group initiated cART only weeks or months prior to 9 months of age, more than half were severely immunosuppressed at initiation, and only 80% reached viral suppression at antibody testing, a reflection of both the timing of the study and the challenges of infant diagnostic testing and treatment.^34–36^ There may not have been sufficient time for immune reconstitution before measles vaccination and thus antibody responses were more similar to CHIV initiating cART after measles vaccination. Studies are needed to determine whether initiating cART within days or weeks of life is effective at preserving measles immunity and efforts should continue to be made to improve timely access to HIV testing and treatment for CHIV.

Antibodies waned quickly among CHIV who were seropositive after 9 months of age, regardless of timing of cART initiation, and after exposure during the outbreak, with an estimated mean time to seroreversion of 2-3 years. This finding is consistent with a prior study from Zambia in which 24% of CHIV measles seropositive at baseline seroreverted over 18 months.^28^ Similarly, only 40% of treatment-naïve CHIV in Zambia were seropositive 27 months after receiving MCV1.^37^

One limitation of this study was the assay sensitivity, which produced lower seroprevalence estimates than expected and necessitated applying a correction factor derived from a second assay. As the correction factor produced results for controls in line with estimates from the national survey in Zambia and a prior study in this setting,^38, 39^ we are confident in the estimates for CHIV and the comparisons between groups. A second limitation was that measles vaccination history was not available for most CHIV and all controls. Given the high vaccine coverage for MCV1 in Southern Province,^17^ we assumed that most CHIV and controls were vaccinated and that lower likelihood of vaccination among CHIV, which has been observed in some but not all studies,^40, 41^ was not responsible for the differences observed. This assumption was supported by the sensitivity analysis which found low seropositivity even among CHIV with documented MCV. The lack of vaccination history also limited our ability to evaluate the impact of immune and viral suppression at vaccination on levels measles antibodies. Immune and viral suppression at cART initiation and measles antibody testing were provided as potential proxies for the late and early cART groups, respectively. A third limitation was that control children were not confirmed HIV uninfected due to restrictions on sample testing and this was addressed by excluding children reported to receive antiretroviral drugs. While it is possible that some were undiagnosed or untreated CHIV, this is unlikely given the high morbidity and mortality experienced by untreated CHIV. A fourth limitation was potential survival bias, particularly among CHIV initiating cART at older ages who may have had slower disease progression. This may account for the higher seropositivity and longer time to seroreversion observed in the older age groups.

In conclusion, low levels of measles IgG antibodies were observed among CHIV younger than 5 years of age, even when cART was initiated before 9 months of age, suggesting that children initiating cART within several months prior to MCV1 remain susceptible to measles would benefit from a second dose of measles-containing vaccine. While most countries have introduced MCV2 between 15 and 18 months of age, coverage is often substantially lower than for MCV1.^42^ For CHIV, MCV2 is critical and they should be prioritized for receipt as part of routine HIV care. If a second dose is given fewer than 6 to 9 months after starting cART, protection is less than optimal and a third dose should be considered, consistent with WHO guidelines.^7^ Lastly, significant waning of measles antibodies was observed in this study after 9 months of age and following boosting in the context of circulating virus and SIAs. Current recommendations for CHIV should be revisited in this era of early cART initiation and routine MCV2, and the need for periodic revaccination throughout childhood and adolescence may need to be considered in settings with a high risk of measles virus infection.^6^

## Supporting information

Supplemental Digital Content

## Acknowledgements

We thank the children and their parents for participating in the studies, and the staff at Macha Research Trust for assisting with the studies.

## Data availability statement

The data used to support the findings of this study are governed by the regulations of the Government of Zambia and are not publicly available. Investigators interested in the data are required to submit a written request to the Ministry of Health. Contact Dr. Catherine Sutcliffe to coordinate the request.

