## Supplemental Digital Content for "Timing of antiretroviral treatment initiation and seropositivity to measles virus among children living with HIV in rural Zambia"

Supplemental Digital Content 1. Study timeline

Supplemental Digital Content 2. PART study participants included in each of the analyses

Supplemental Digital Content 3. Measles IgG Antibody Testing and Quantitative Adjustment

Supplemental Digital Content 4. Evaluation of the impact of sample type and storage duration on measles antibody concentrations

Supplemental Digital Content 5. Sensitivity analysis restricted to PART study participants with available data on measles vaccination status

Supplemental Digital Content 6. Distribution of age in years at the time of first serology sample by timing of treatment initiation.

Supplemental Digital Content 7. Distribution of age at receipt of first dose of measles-containing vaccine (MCV1) by timing of treatment initiation.

Supplemental Digital Content 8. Number of individuals and time points included in decay analysis

Supplemental Digital Content 9. Longitudinal analysis methods

Supplemental Digital Content 10. Sensitivity analysis of the post-MCV1 antibody decay analysis changing the criteria for censoring

Supplemental Digital Content 11. Sensitivity analysis of the post-MCV1 antibody decay analysis including all children living with HIV with at least one observation to contribute to the analysis

Supplemental Digital Content 12. Correlates of seropositivity among children living with HIV

Supplemental Digital Content 13. Measles antibody seroprevalence by HIV status and calendar year among children included in the cross-sectional analysis

Supplemental Digital Content 14. Complete individual-level trajectories from the 195 individuals who had at least one observed boost during follow-up

Supplemental Digital Content 15. Individuals with observed boosting episodes during follow-up and probability of observed boosting

Supplemental Digital Content 16. Changes in measles antibody concentration during the 2010-2011 outbreak period

### Supplemental Digital Content 1. Study timeline

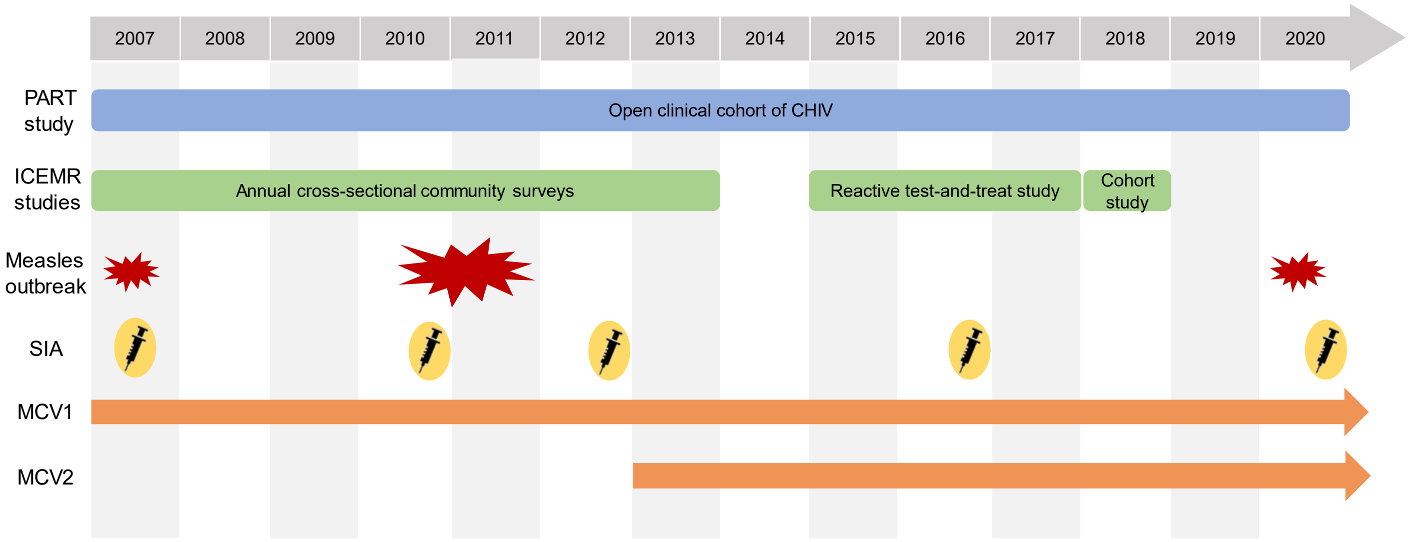


ICEMR: International Centers of Excellence for Malaria Research; MCV: measles-containing vaccine; PART: Pediatric Antiretroviral Treatment; SIA: supplementary immunization activity

### Supplemental Digital Content 2. PART study participants included in each of the analyses

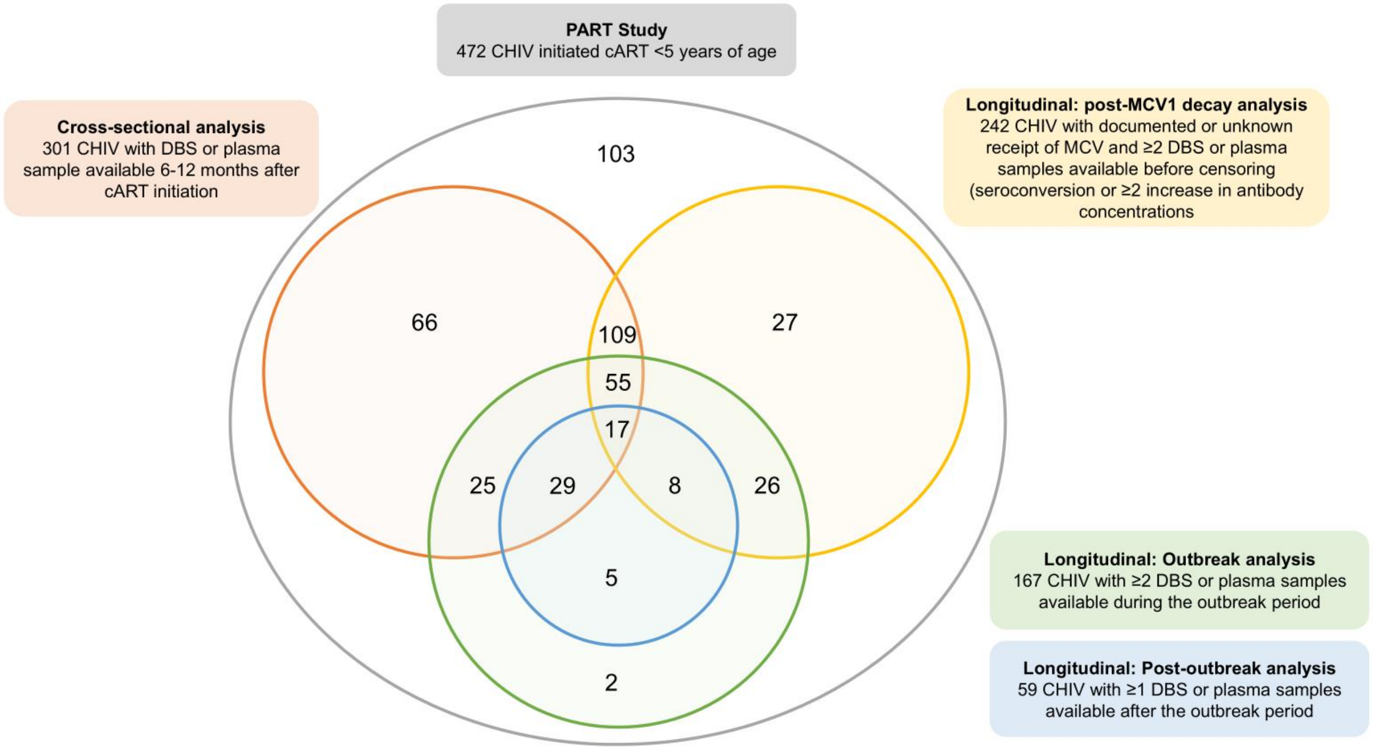


DBS: dried blood spot; MCV: measles-containing vaccine; PART: Pediatric Antiretroviral Treatment

**Supplemental Digital Content 3. Measles IgG Antibody Testing and Quantitative Adjustment**

**Measles IgG Antibody Testing – Additional Information**

DBS and plasma samples were tested for anti-measles virus IgG antibodies in the Clinical Research Laboratory at Macha Research Trust using the Euroimmun Anti-Measles Virus ELISA (Euroimmun AG, Lübeck, Germany), which was found to have a sensitivity and specificity of 100% each when tested against a panel of 112 clinical characterized specimens.[1] Samples for this study were tested according to manufacturer recommendations. Four calibrators provided were run with the samples and a standard curve was calculated and used to convert optical density (OD) values to mIU/mL. Based on a prior study,[2] results between 100-149 mIU/mL were considered equivocal and retested, with repeat results considered final. Samples with results above the highest calibrator (5000 mIU/mL) were retested at a dilution of 1:4, as recommended by the manufacturer. Results below the lower limit of detection of the assay (8 mIU/mL) were assigned a value of 8 mIU/mL.

**Measles IgG Antibody Quantitative Adjustment**

*Rationale*

Preliminary results from this study and a serosurvey conducted in Zambia in 2020 yielded much lower than anticipated estimates of measles IgG seropositivity, given estimated vaccine coverage and results from prior studies conducted in this setting.[3,4] In addition, prior issues identified by the research team with the Euroimmun measles enzyme immunoassay (EIA) (Euroimmun AG, Lübeck, Germany) after a change was made to one of the calibrators that primarily impacted specimens near the equivocal threshold added to concerns about the low sensitivity of this assay.[2] To evaluate the performance of the assay, a comparative study was conducted.

*Source of samples*

In November 2020, a measles serosurvey was nested within a measles supplemental immunization activity (SIA) in Zambia.[5] The serosurvey was conducted among children aged 9 months to 5 years enrolled at selected vaccination sites, including static sites at health centers and outreach sites, in Choma District, Southern Province and Ndola District, Copperbelt Province. Blood was collected by finger prick and spotted onto Whatman 903 filter paper by trained survey staff. Dried blood spot (DBS) cards were dried, processed and then stored at -20°C at the Tropical Diseases Research Centre (TDRC) in Ndola District and Macha Research Trust (MRT) in Choma District.

*Measles EIA testing*

DBS were eluted and tested for anti-measles virus IgG antibodies at the TDRC and MRT laboratories. Samples were tested using the Euroimmun measles EIA (Euroimmun AG, Lübeck, Germany) according to manufacturer recommendations. Values $\geq$ 200 mIU/mL were assigned positive, 150 - 199 equivocal and <150 negative. Equivocal samples were retested and the repeat result was treated as final. Samples above the top calibrator (5000 mIU/mL) were not retested. Samples below the bottom calibrator were set to the lower limit of detection (8 mIU/mL). Four randomly selected samples were run in duplicate to monitor intra-plate variability. Every 20$^{th}$ specimen per plate was retested to assess inter-plate variability.

*Sample selection for the comparative study*

To assess the sensitivity of the EIA, a subsample of 300 DBS specimens were selected from those collected in Ndola District during the serosurvey. The specimens were purposefully selected across the range of values, with more samples selected with values around the thresholds. Half of the specimens had EIA antibody values between 100-200 mIU/mL, 100 had values <100 mIU/mL, and 50 had values >200 mIU/mL. Samples were shipped to the Centers for Disease Control and Prevention (CDC), Viral Preventive Diseases Branch in Atlanta, Georgia, USA for testing using a multiplex bead assay (MBA).

*Measles MBA testing*

The MBA is a serological assay that has been increasingly utilized for its ability to provide high-quality seroprevalence data and to test antigens from multiple pathogens at once,[6] among other factors. The MBA uses a commercially produced whole-virus lysate antigen prepared using MagPlex or MicroPlex beads. In a validation study comparing the MBA to the measles plaque reduction neutralization (PRN) assay, the current "gold standard", there was a strong correlation between IgG concentrations from the MBA and PRN titers ($R = 0.846$).[6] Compared to PRN, the MBA had a sensitivity of 98% and a specificity of 83%, when a seroprotection cutoff at 153 mIU/mL was used.[5]

In the comparative study, samples were tested using a MagPix instrument (Luminex Corp., Austin, Texas), and including background beads with uninfected Vero lysate to account for background signals in the specimens. All plates were run with a standard curve, which was used to calculate observed concentrations.

*Statistical analysis*

For the samples tested using both the EIA and MBA, results from both assays were compared. There appeared to be a linear relationship between the two assay outputs, with the MBA producing consistently higher results than the EIA (Figure S1), particularly at higher concentrations (Figure S2). As this supported a low sensitivity of the Euroimmun assay, an analysis was performed to estimate a correction factor to be applied to the EIA IgG antibody concentrations. We fit models to a subset of data between 8 mIU/mL EIA (the lower limit of detection of the EIA) and 3000 mIU/mL EIA (removing the outlier) (Figure S1). We employed the 'brms' package, which is a Bayesian statistical package in R, to fit two linear models to the data. We found the log-log linear model fit the data best compared to linear model (Figure S3). Figure S4 displays a graphical representation of the log-log model fit to the data. The mean estimate of the intercept was 0.495 and the mean estimate of the slope was 1.092, yielding the following equation as a correction factor to calculate adjusted mIU/mL: Adjusted_mIUmL = exp(0.495 + (1.092*ln(EIA_mIUmL)).

The correction factor was applied to the entire range of EIA values from the cross-sectional and longitudinal analyses in this study. Adjusted values at or above 153 mIU/mL were considered seropositive. After applying the correction factor to the EIA values from the cross-sectional analysis, the proportion seropositive increased from 27% to 40% among children living with HIV and from 61% to 77% among controls (Figure S5). The proportions seropositive at a threshold of 120 mIU/mL are also provided for comparison.

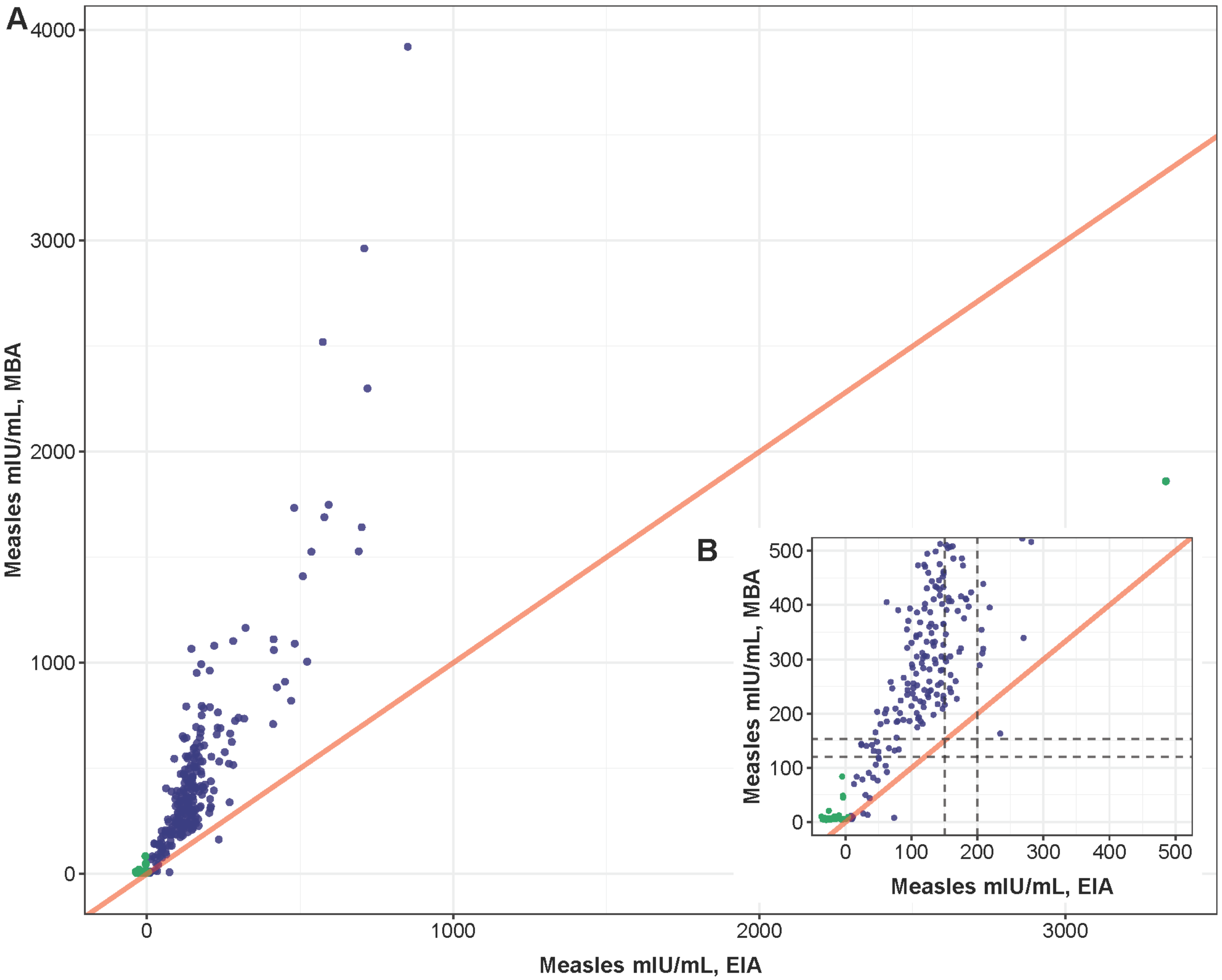


**SDC 3: Figure S1. Scatter plot comparing IgG antibody concentrations from the MBA and EIA (N=290).** A) Scatter plot of the full range of data and B) scatter plot of data less than 500 mIU/mL. The red line represents perfect agreement between the two tests. The green points represent specimens that per the EIA are >3000 mIU/mL or <8 mIU/mL, which were removed prior to fitting the linear models. The dashed line on the y-axis indicates the MBA cutoff for seroprotection of 153 mIU/mL and 120 mIU/mL. The dashed lines on the x-axis indicate the thresholds for borderline (150 mIU/mL) and positive (200 mIU/mL) in assessing immune status using the EIA.

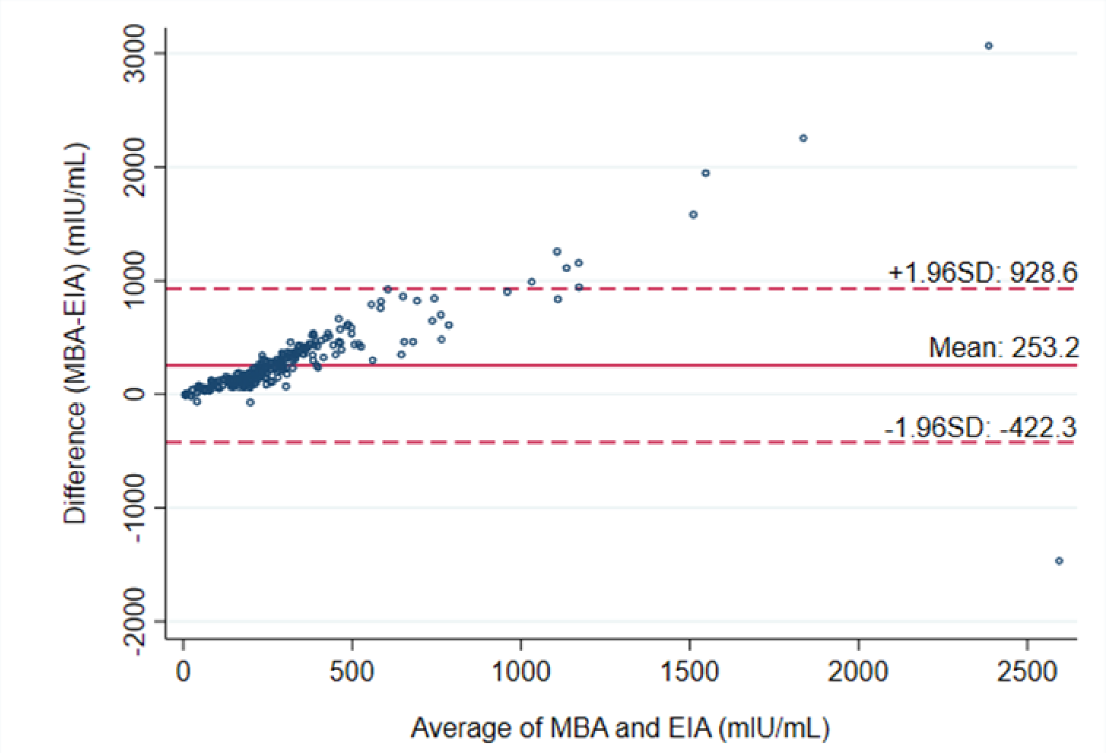


**SDC 3: Figure S2. Bland-Altman plot comparing the difference in IgG antibody concentrations from the MBA and EIA, relative to their average antibody concentration.** The solid red line indicates the average difference between MBA and EIA IgG antibody concentrations, while the dashed red lines indicate thresholds for differences between MBA and EIA concentrations that are ± 1.96 standard deviations (SD) from the mean.

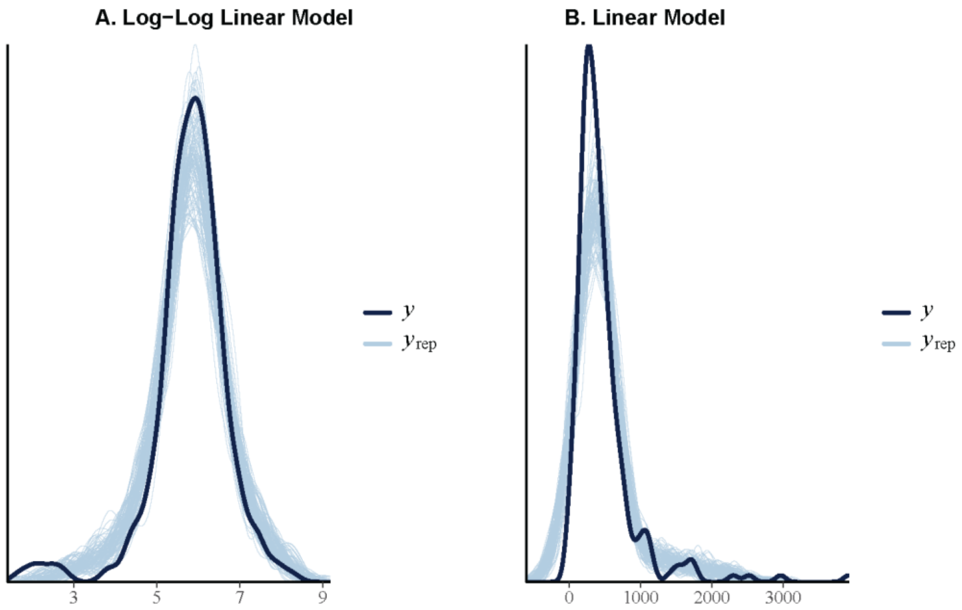


**SDC 3: Figure S3. Distribution of observed and replicated measles IgG antibody concentrations (mIU/mL).** Posterior predictive checks of Log-Log Linear Model (A) and Linear Model (B). The dark line (y) represents the observed MBA data, and the light blue lines ($y_{rep}$) represent predictions of MBA from the posterior distributions. We see that the log-log model fit the data better.

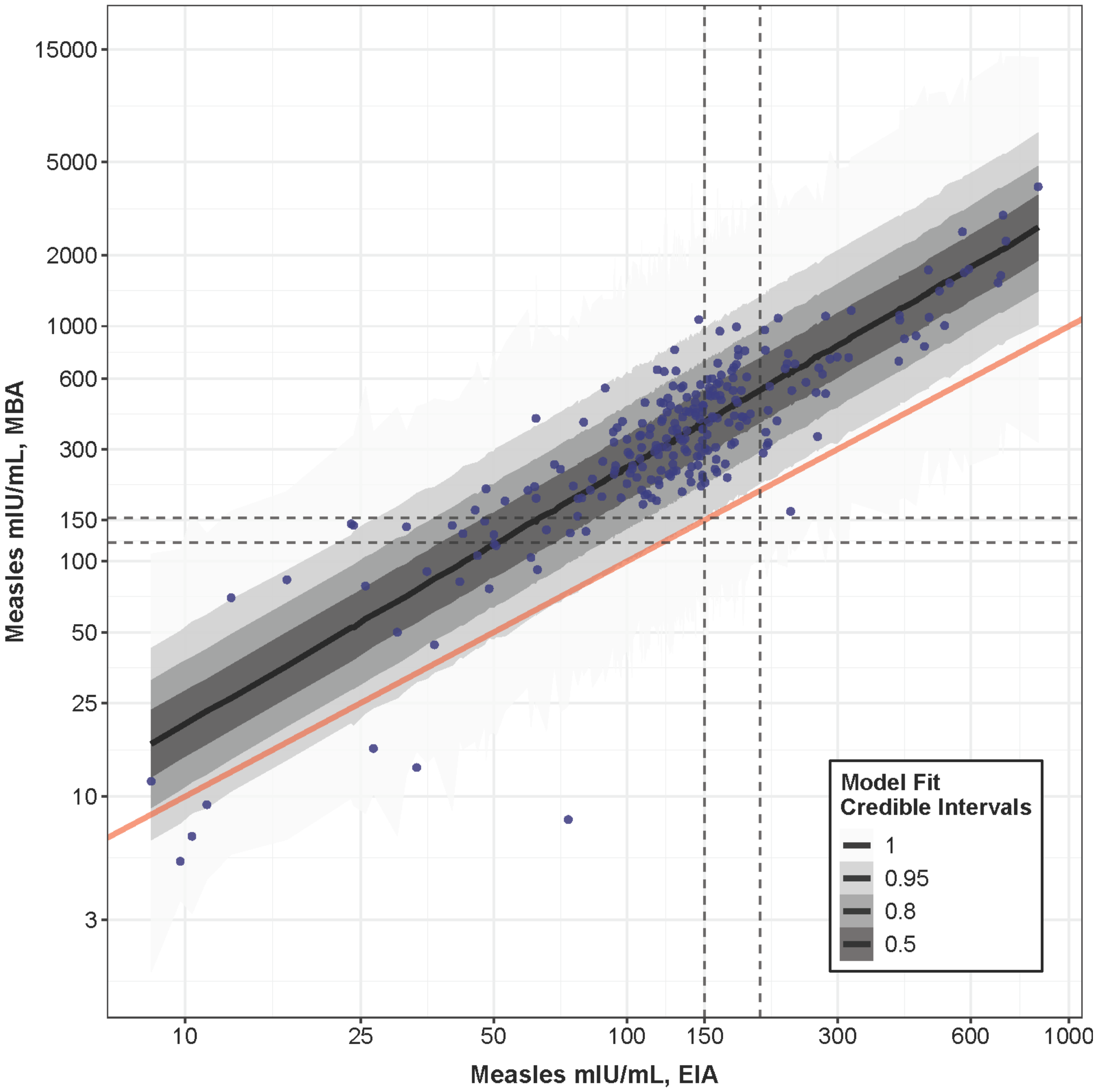


**SDC 3: Figure S4. Log-log model fit for EIA versus MBA (N=249).** The blue points represent the data. The black line indicates the mean fitted regression line, while the red line indicates a reference line of equivalence between the EIA and MBA. The ribbons of different shades of grey represent the 50%, 80%, 95%, and 100% credible intervals. The dashed line on the y-axis indicates the MBA cutoff for seroprotection of 153 mIU/mL and 120 mIU/mL. The dashed lines on the x-axis indicate the thresholds for borderline (150 mIU/mL) and positive (200 mIU/mL) in assessing immune status using the EIA.

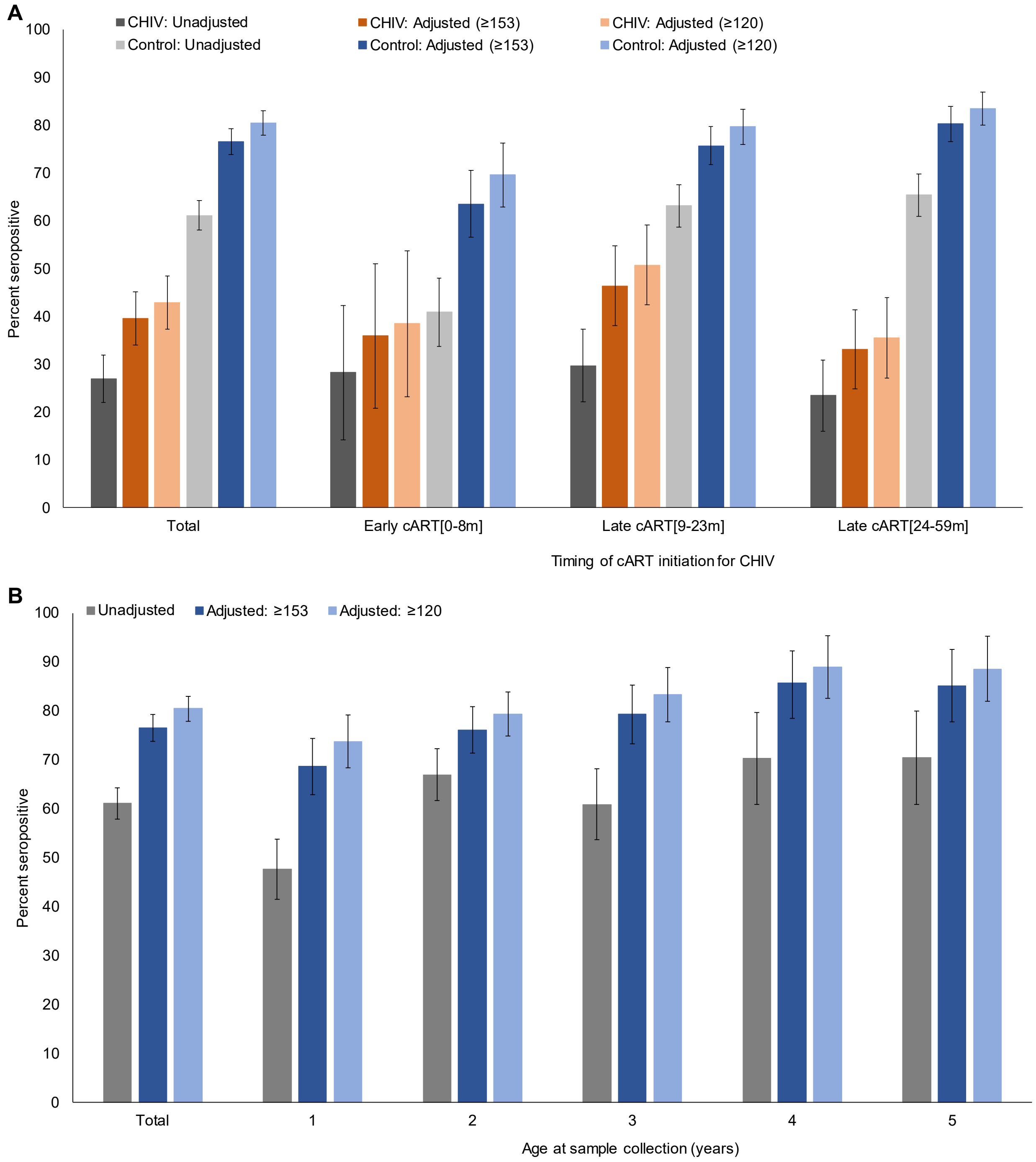


**SDC 3: Figure S5. Cross-sectional analysis of measles seropositivity before and after applying the correction factor for (A) children living with HIV (CHIV) by timing of ART initiation and age-matched HIV-uninfected controls and (B) HIV-uninfected children by age at sample collection.** For CHIV, results are provided overall and stratified by age of treatment initiation relative to expected age of primary measles vaccination at 9 months: Early cART[0-8m], Late cART[9-23m], and Late cART[24-59m]. HIV-uninfected controls were age-matched to CHIV based on age at measles antibody testing which occurred 6-12 months after treatment initiation. The unadjusted results represent the IgG antibody concentrations obtained from the EIA; values above 150 mIU/mL were considered seropositive (i.e. borderline values of 150-199 mIU/mL were considered positive). The adjusted results represent the IgG antibody concentrations obtained from the EIA with the correction factor applied; adjusted values ≥153 mIU/mL were considered seropositive for the main analysis; adjusted values ≥120 mIU/mL are also provided for comparison. Error bars represent 95% confidence intervals.

**Supplemental Digital Content 4. Evaluation of the impact of sample type and storage duration on measles antibody concentrations**

*Rationale*

This study included stored samples collected from children between 2007 and 2020 who were enrolled in two studies in rural Zambia – the Pediatric ART study (PART) and International Center for Excellence in Malaria Research (ICEMR) studies. While stored samples from the ICEMR studies were all dried blood spots (DBS), samples from the PART study included both DBS and plasma samples. To determine the potential impact of sample type and storage duration on measles antibody concentrations, we conducted a comparative study using paired DBS and plasma samples across the range of years observed in the main study.

*Source of samples*

Samples were selected from two sources:

1) The PART study, a clinical cohort study of children living with HIV (CHIV) conducted at the Macha HIV clinic in Choma District, Southern Province, Zambia from 2007 to 2023. At study visits, twice a year a venous blood sample was collected from the child and stored as frozen plasma at -80°C and a DBS card at -20°C in the Clinical Research Laboratory at Macha Research Trust.

2) Venous blood samples were collected from three community volunteers in 2022 to serve as internal controls for the testing. Samples were also stored as frozen plasma at -80°C and a DBS card at -20°C in the Clinical Research Laboratory at Macha Research Trust.

*Sample selection for comparative study*

Samples from the PART study were eligible if both a DBS card and plasma sample were available in sufficient quantity for testing and if they were collected between 2010 and 2018. Samples from 2010 and 2012 were eligible if they were tested in the prior measles study that was nested in the PART study among adolescent CHIV.[1] Samples were purposively selected to cover the range of values (negative, equivocal, low positive, medium positive, and high positive) of IgG antibody concentrations measured using the Enzygnost enzyme immunoassay (Siemens, Munich, Germany). A convenience sample of paired samples from 2014, 2016, and 2018 were selected. In addition, paired plasma and DBS samples from the internal controls were selected.

*Measles EIA testing*

DBS and plasma samples were tested for anti-measles virus IgG antibodies in the Clinical Research Laboratory at Macha Research Trust. Samples were tested using the Euroimmun Anti-Measles Virus ELISA (Euroimmun AG, Lübeck, Germany) according to manufacturer recommendations. DBS (6mm) were eluted according to manufacturer instructions. Paired DBS and plasma samples collected from each of the years as well as at least one pair from each of the internal controls were run on each plate. Samples below the bottom calibrator were set to the lower limit of detection (8 mIU/mL).

*Statistical analysis and results*

For the paired samples of DBS and plasma, results from both sample types were compared (Figure S1 and S2). There appeared to be a linear relationship between the two assay outputs, with plasma samples producing consistently higher results than the DBS (Figure S1), particularly at higher concentrations (Figure S2). An analysis was performed to estimate a correction factor to be applied to the plasma IgG antibody concentrations. We elected to apply a correction factor to the plasma rather than the DBS IgG antibody concentrations, as all samples for control children in the cross-sectional analysis were DBS while samples for CHIV were a mix of plasma and DBS samples. For consistency in sample types across groups, the correction factor was applied to the plasma samples from CHIV. The same correction factor was applied to the plasma samples for the longitudinal analysis for consistency across analyses.

We fit models to the data by employing the 'brms' package, which is a Bayesian statistical package in R, to fit a log-log linear model without an intercept (Figure S1). Figure S3 displays a graphical representation of the log-log model fit to the data. The mean estimate of the slope was 0.84447, yielding the following equation as a correction factor to calculate adjusted mIU/mL for plasma samples: AdjustedDBS_mIUmL = exp(0+(0.84447*ln(plasma_mIUmL))).

There did not appear to be a relationship between IgG antibody concentrations across year for DBS or plasma samples (Figure S4 and S5).

The correction factor was applied to the entire range of EIA values from plasma samples included in the cross-sectional (167 of 301 samples; 55%) and longitudinal analyses (1,059 of 1,818 samples; 58%) from the PART study. Adjusted values <8 mIU/mL were set to 8 mIU/mL. The second correction factor from the MBA validation study (see Supplemental Digital Content 3) was then applied, with any resulting values at or above 153 mIU/mL considered positive. After applying the two correction factors to the EIA values in the cross-sectional analysis, the proportion seropositive among CHIV increased from 27% to 32%, but was lower than the proportion seropositive with only the second correction factor applied from the MBA validation study (40%; Figure S6).

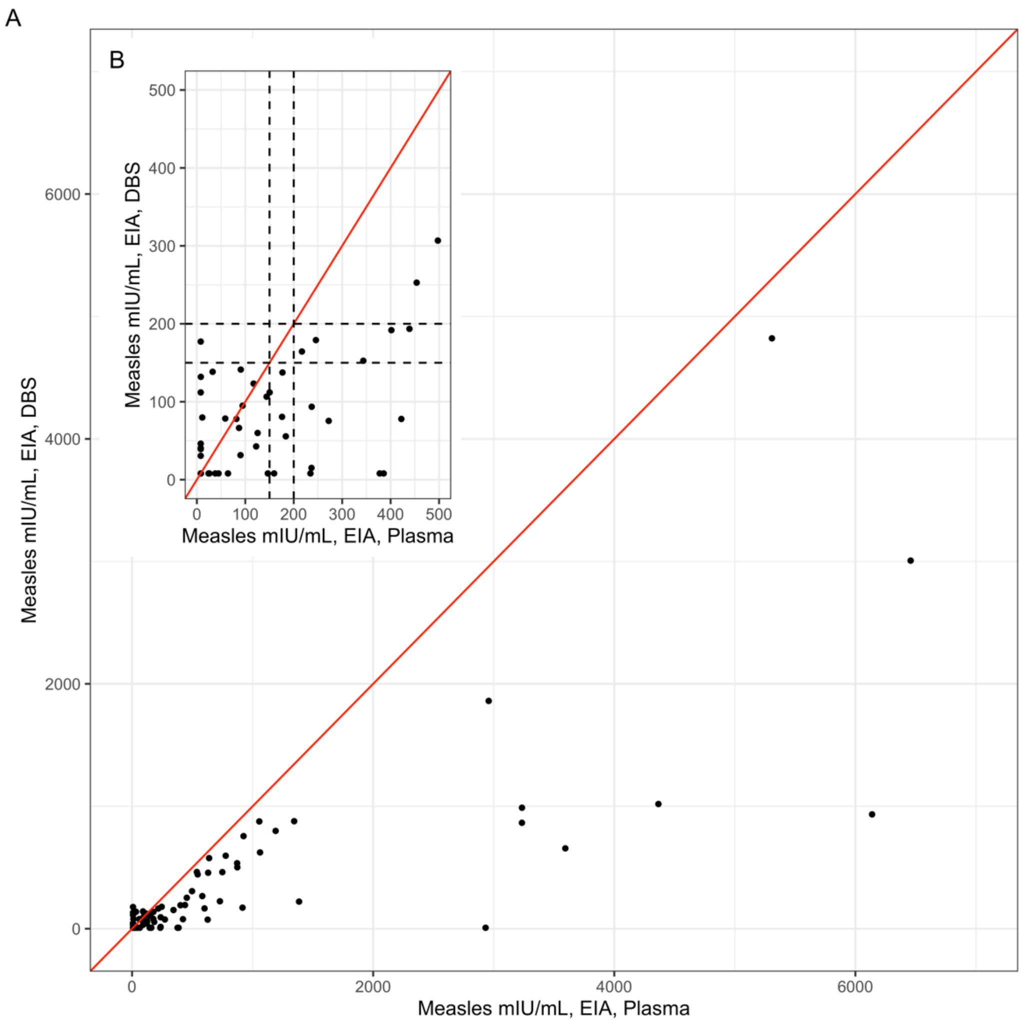


**SDC 4: Figure S1. Scatter plot comparing IgG antibody concentrations from paired plasma and DBS samples tested using the EIA (N=74).** A) Scatter plot of the full range of data and B) scatter plot of data less than 500 mIU/mL for both sample types. The red line represents perfect agreement between the two tests. The dashed lines on the x-axis and y-axis indicate the thresholds for borderline (150 mIU/mL) and positive (200 mIU/mL) in assessing immune status using the EIA.

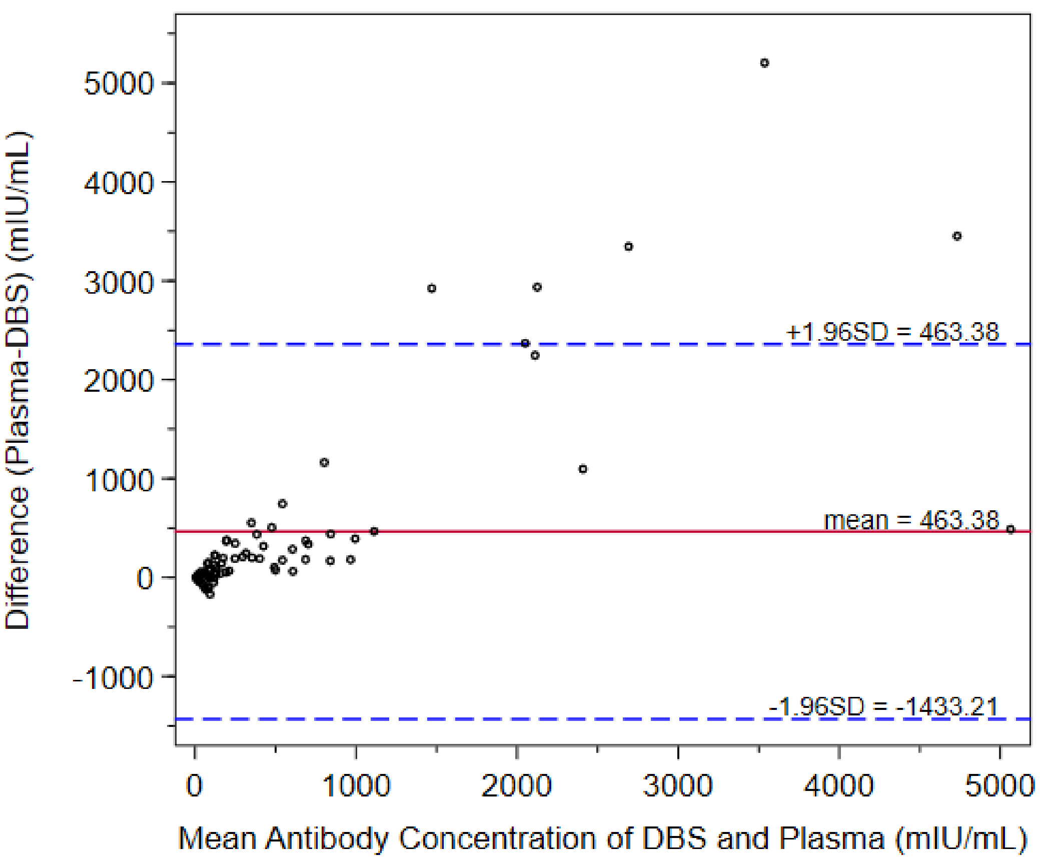


**SDC 4: Figure S2. Bland-Altman plot comparing the difference in IgG antibody concentrations from paired plasma and DBS samples, relative to their average antibody concentration.** The red line indicates the mean of differences between DBS and plasma antibody concentrations, while the dashed blue line indicates ± 1.96 standard deviations from the mean.

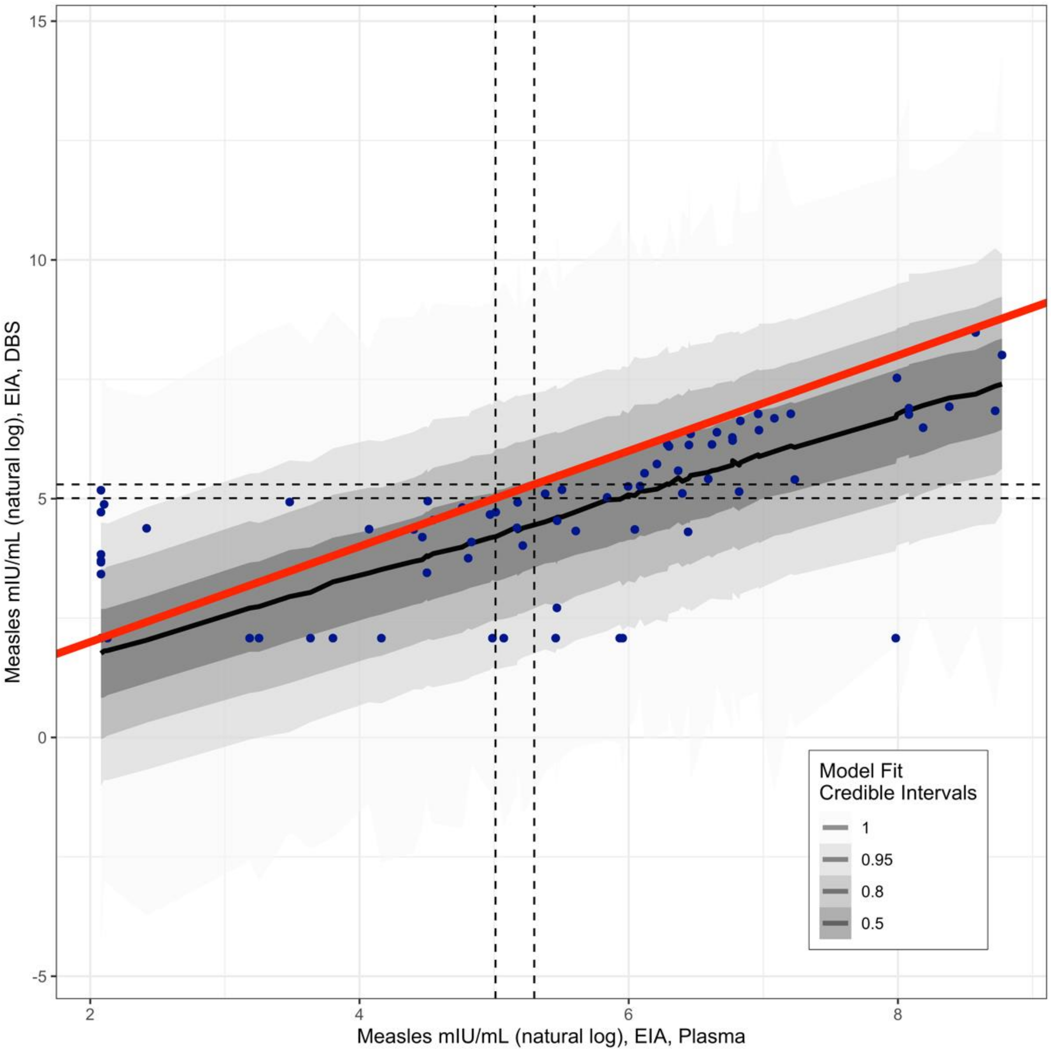


**SDC 4: Figure S3. Log-log model fit for plasma versus DBS (N=74).** The blue points represent the data. The black line indicates the mean fitted regression line, while the red line indicates a reference line of equivalence between the paired plasma and DBS samples. The ribbons of different shades of grey represent the 50%, 80%, 95%, and 100% credible intervals. The dashed lines on the x-axis and y-axis indicate the thresholds for borderline (150 mIU/mL) and positive (200 mIU/mL) in assessing immune status using the EIA.

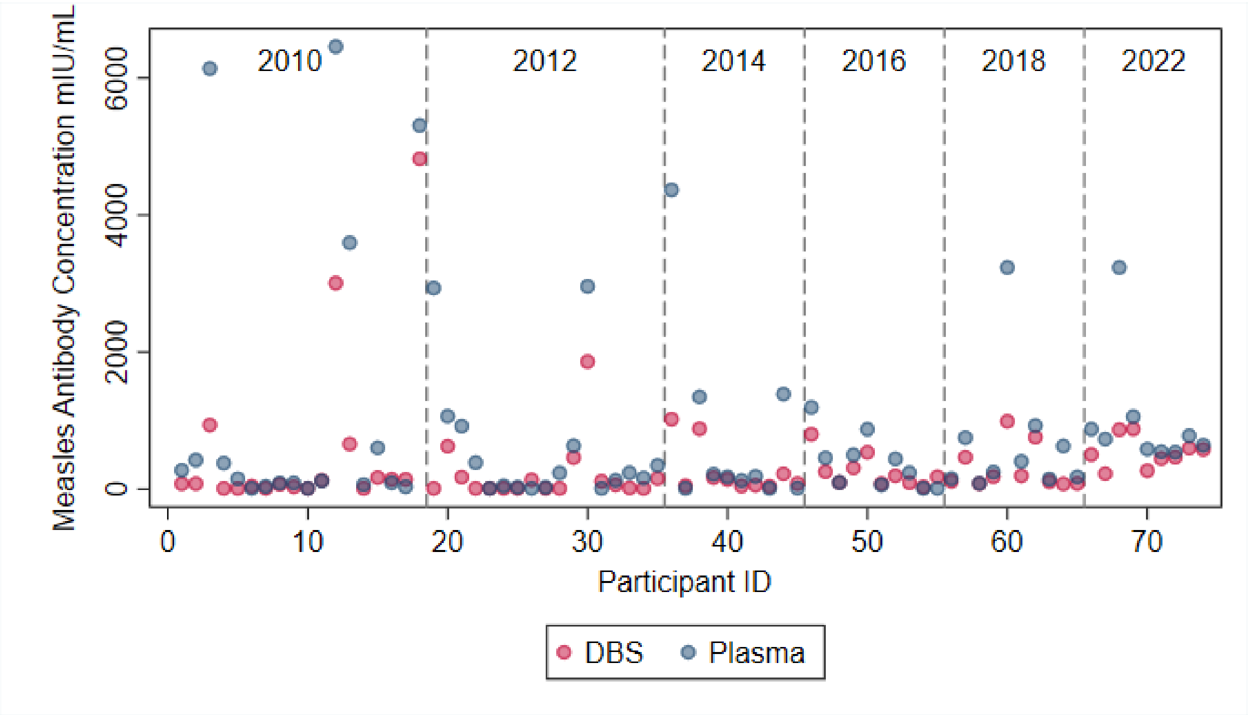


**SDC 4: Figure S4. Dried blood spot (DBS) and plasma IgG antibody concentrations of paired samples by sample ID and year of collection.**

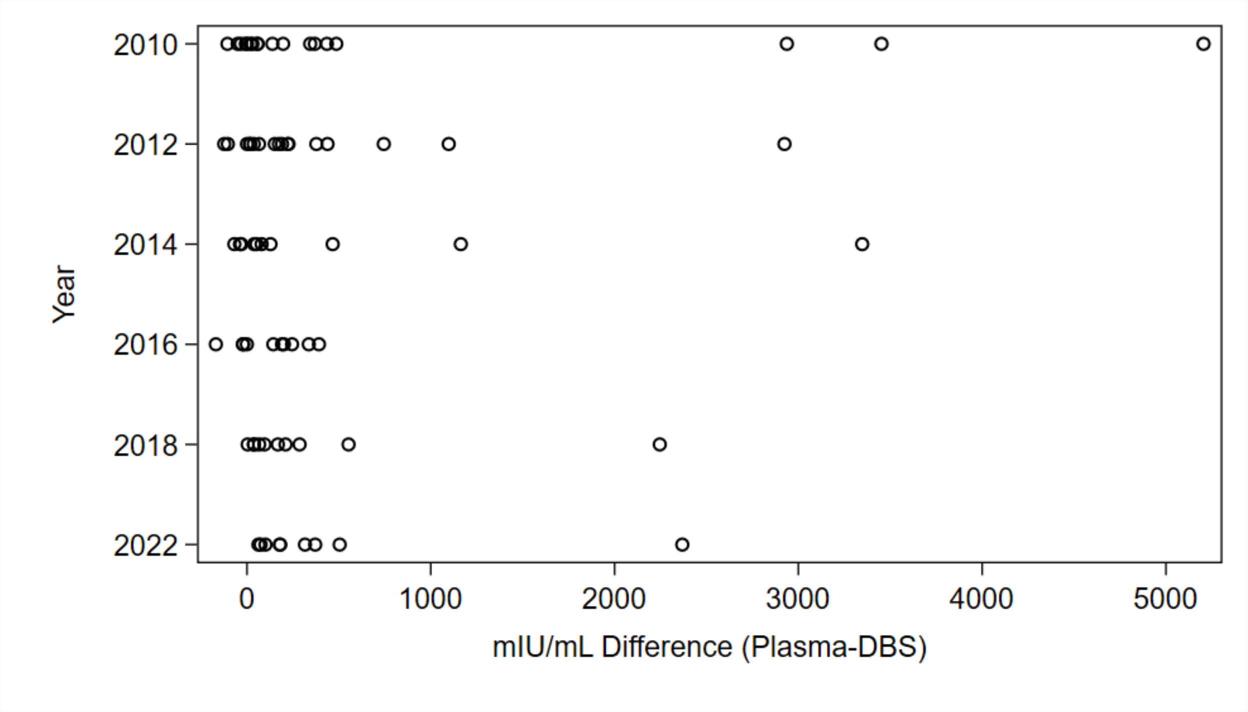


**SDC 4: Figure S5. Difference between dried blood spot (DBS) and plasma IgG antibody concentrations in paired samples by year of collection.**

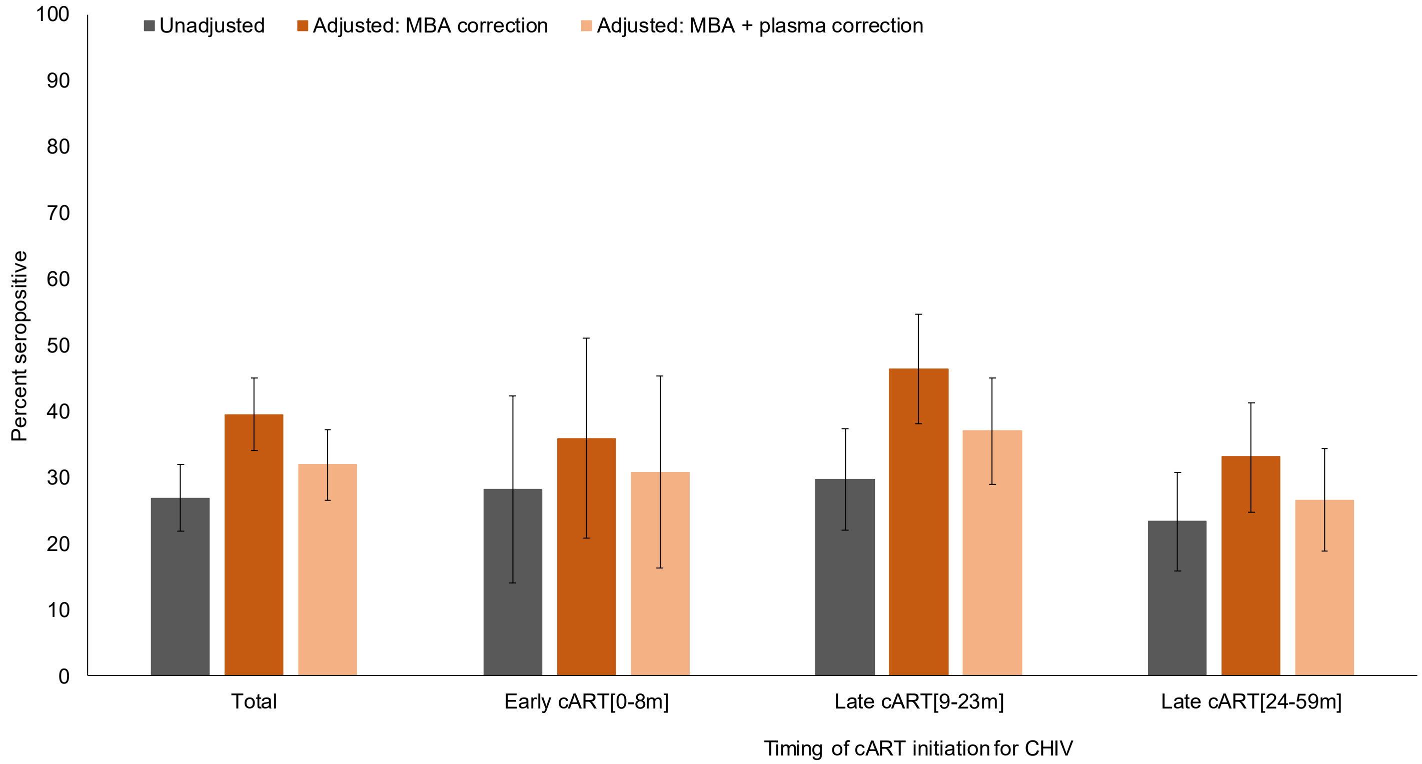


**SDC 4: Figure S6. Cross-sectional analysis of measles seropositivity among children living with HIV (CHIV) before and after applying the correction factors.** For CHIV, results are provided overall and stratified by age of treatment initiation relative to expected age of primary measles vaccination at 9 months: Early cART[0-8m], Late cART[9-23m], and Late cART[24-59m]. The unadjusted results represent the IgG antibody concentrations obtained from the EIA; values ≥150 mIU/mL were considered seropositive (i.e. borderline values of 150-199 mIU/mL were considered positive). The adjusted results with the multiplex bead assay (MBA) correction represent the IgG antibody concentrations obtained from the EIA with a correction factor applied from the MBA validation study (see Section 1); adjusted values ≥153 mIU/mL were considered seropositive. The adjusted results with the MBA and plasma correction represent the IgG antibody concentrations obtained from the EIA with the correction factor applied to plasma samples as described above and the MBA correction factor applied to all samples; adjusted values ≥153 mIU/mL were considered seropositive. Error bars represent 95% confidence intervals.

**Supplemental Digital Content 5. Sensitivity analysis restricted to PART study participants with available data on measles vaccination status**

Of the 301 children living with HIV (CHIV) included in the cross-sectional analysis, 57 (29%) had data available on measles vaccination status, including 1 (3%) in the Early cART[0-8m] group, 26 (19%) in the Late cART[9-23m] group, and 33 (27%) in the Late cART[24-59m] group. Among the 57 CHIV with available data (median age: 3.0 years; interquartile range: 2.3, 4.1), 53 (93%) had documented receipt of a measles-containing vaccine at a median age of 9.9 months (interquartile range: 8.9, 11.1). No statistically significant differences were found in measles antibody seroprevalence between those with and without data available on measles vaccine status.

**Measles antibody seroprevalence among children living with HIV (CHIV) by measles vaccine status and timing of treatment initiation**

| | Total | | CHIV with measles vaccine data | | CHIV who received measles vaccine | | CHIV without measles vaccine data | |
|---|---|---|---|---|---|---|---|---|
| | N | n (%) | N | n (%) | N | n (%) | N | n (%) |
| Overall | 301 | 96 (31.9) | 57 | 21 (36.8) | 53 | 20 (37.7) | 244 | 75 (30.7) |
| Early cART[0-8m] | 39 | 12 (30.8) | 1 | 1 (100.0) | 1 | 1 (100.0) | 38 | 11 (29.0) |
| Late cART[9-23m] | 138 | 51 (37.0) | 26 | 13 (50.0) | 24 | 12 (50.0) | 112 | 38 (33.9) |
| Late cART[24-59m] | 124 | 33 (26.6) | 30 | 7 (23.3) | 28 | 7 (25.0) | 94 | 26 (27.7) |

**Supplemental Digital Content 6. Distribution of age in years at the time of first serology sample by timing of treatment initiation**

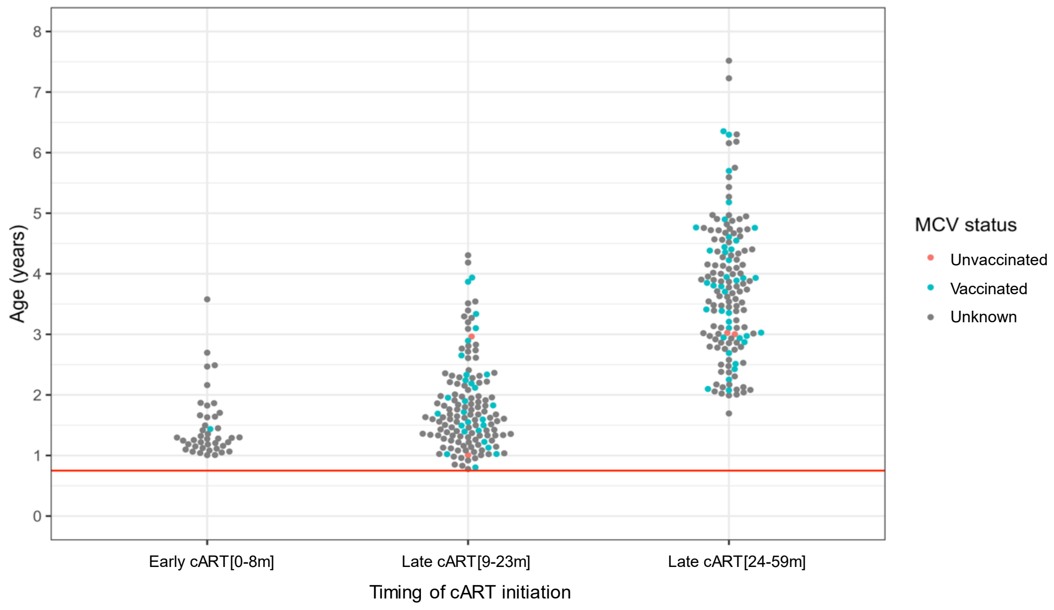


Results are stratified by age of treatment initiation relative to expected age of primary measles vaccination at 9 months: Early cART[0-8m], Late cART[9-23m], and Late cART[24-59m]. The color of the point represents whether individuals were known to be unvaccinated (red, n=4), the date of MCV receipt was recorded (blue, n=66), or assumed to have occurred exactly at 9 months of age (grey, n=264). The red horizontal line indicates 9 months of age.

**Supplemental Digital Content 7. Distribution of age at receipt of first dose of measles-containing vaccine (MCV1) by timing of treatment initiation**

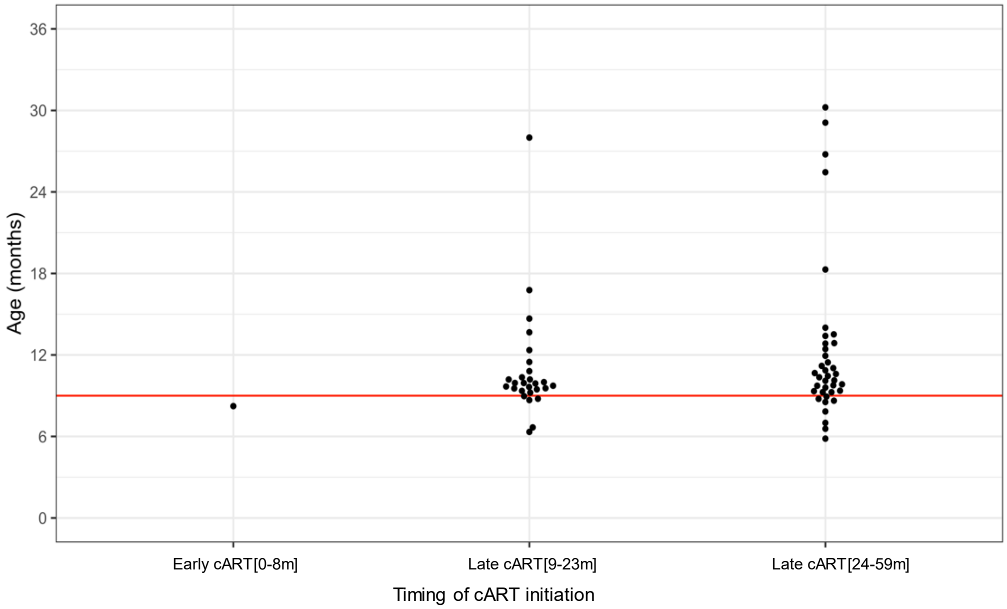


Results for the 66 participants with documented age at MCV1 are stratified by age of treatment initiation relative to expected age of primary measles vaccination at 9 months: Early cART[0-8m], Late cART[9-23m], and Late cART[24-59m]. The horizontal red line indicates 9 months of age.

**Supplemental Digital Content 8. Number of individuals and time points included in decay analysis**

| | Early cART [0-8m] | Late cART [9-23m] | Late cART [24-59m] | Total |
|---|---|---|---|---|
| **Number of participants** | **42** | **141** | **147** | **330** |
| **MCV1 date available** | **1** | **27** | **38** | **66** |
| 1 time point | 0 | 11 | 8 | 19 |
| 2 time points | 0 | 4 | 13 | 17 |
| 3 time points | 0 | 2 | 10 | 12 |
| 4 time points | 1 | 6 | 4 | 11 |
| 5 time points | 0 | 2 | 1 | 3 |
| 6 time points | 0 | 2 | 0 | 2 |
| 7 time points | 0 | 0 | 1 | 1 |
| 8 time points | 0 | 0 | 1 | 1 |
| **MCV1 date not available** | **41** | **114** | **109** | **264** |
| 1 time point | 11 | 30 | 28 | 69 |
| 2 time points | 12 | 23 | 25 | 60 |
| 3 time points | 12 | 18 | 17 | 47 |
| 4 time points | 2 | 20 | 11 | 33 |
| 5 time points | 4 | 10 | 13 | 27 |
| 6 time points | 0 | 9 | 7 | 16 |
| 7 time points | 0 | 4 | 5 | 9 |
| 8 time points | 0 | 0 | 2 | 2 |
| 9 time points | 0 | 0 | 1 | 1 |

cART: combination antiretroviral therapy; MCV: measles-containing vaccine.

**Supplemental Digital Content 9. Longitudinal analysis methods**

*Model structure for Bayesian growth (mixed effects) mixture model: initial post-MCV1 decay*

$Y_{ijk}$ is the log adjusted titer for individual *i*, time *j*, in cART group *k* = {1,2,3}, and $X_{ijk}$ is years since MCV1. We assumed that each individual is from one of two latent serological classes:

1. If individual *i* belongs to **serological class 1** ("non-responders"):

   $Y_{ijk} \sim \text{Normal}(\text{LOD}, \sigma^2)$, truncated at the lower limit of detection (LOD)

2. If individual *i* belongs to **serological class 2** ("decayers"):

   $Y_{ijk} \sim \text{Normal}((\alpha_k+\delta_i)+\beta_k * X_{ijk}, \sigma^2)$, again truncated at the LOD, where:
   - $\alpha_k$ is an cART group-specific intercept (i.e., log antibody concentration at MCV1 receipt, assuming no lag between MCV1 and the peak concentration)
   - $\delta_i$ is a random intercept for individual *i*
   - $\beta_k$ is an cART group-specific slope (i.e., decay of log antibody concentrations as a function of years since receiving MCV1)
   - $\delta_i \sim \text{Normal}(0, \tau^2)$

We assumed that each individual in cART group *k* has a probability of belonging to serological class 1, $w_k$ (and a probability of belonging to serological class 2, $1-w_k$). Weakly informative priors of $\text{Normal}(0, 10)$ were set for $\alpha_k$ and $\beta_k$. We estimated all parameters ($w_k$, $\alpha_k$, $\beta_k$, $\tau^2$, $\sigma^2$) using the Stan programming language (Stan Development Team, https://mc-stan.org/). Using these estimates, we simulated 1,000 potential longitudinal trajectories for each ART group, and subsetted them to trajectories where $\exp(Y_{ijk})$ was greater than 153 mIU/mL at $X_{ijk}=0$, the time of MCV1 receipt. Among these trajectories, we estimated $T_k$, the cART group-specific mean time to seroreversion after MCV1 vaccination specifically among decayers who are seropositive at the time of MCV receipt, as: $T_k=(\text{cutoff}-\alpha_k)/\beta_k$, using a cutoff of 153 mIU/mL on the log scale.

To test the effects of our inclusion criteria for the decay analysis, we performed sensitivity analyses where individuals were censored at sero-conversion or at ≥4-fold increase in antibody titer and included those individuals with only 1 observed time point. The overall findings did not change in these sensitivity analyses.

*Model structure for mixed effects model: post-observed seroconversion*

$Y_{ij}$ is the log adjusted titer for individual *i*, time *j*, and $X_{ij}$ is years since end of the outbreak period, which we defined to be August 1, 2011. We fit a simple model of log-linear decay of antibody titers over time:

$Y_{ij} \sim \text{Normal}((\alpha+\delta_i)+\beta * X_{ij}, \sigma^2)$, where:
- $\alpha$ is an overall intercept
- $\delta_i$ is a random intercept for individual *i*
- $\beta$ is an overall slope
- $\delta_i \sim \text{Normal}(0, \tau^2)$

Using these estimates, we simulated 1,000 potential longitudinal trajectories, and subsetted them to trajectories where $\exp(Y_{ij})$ was greater than 153 mIU/mL at $X_{ij}=0$, the approximate time of measles seroconversion. Among these trajectories, we estimated $T$, the mean time to seroreversion specifically among individuals whose trajectory starts as seropositive, as: $T=(\text{cutoff}-\alpha/\beta)$, using a cutoff of 153 mIU/mL on the log scale.

**Supplemental Digital Content 10. Sensitivity analysis of the post-MCV1 antibody decay analysis changing the criteria for censoring**

| cART group (k) | Probability of being a non-responder ($w_k$) | Probability of being a decayer ($1-w_k$) | ART group-specific intercept, among decayers ($\alpha_k$) | ART group-specific slope, among decayers ($\beta_k$) | Mean time to seroreversion, among decayers who are seropositive at the time of MCV1 receipt, years ($T_k$) | Proportion of decayers who are seropositive at the time of MCV1 receipt |
|---|---|---|---|---|---|---|
| **Early cART[0-8m]** | 0.53 (0.35-0.70) | 0.47 (0.30-0.65) | 6.31 (5.60-7.07) | -0.72 (-0.96, -0.48) | 2.39 (1.79-3.41) | 0.83 (0.66-0.94) |
| **Late cART[9-23m]** | 0.41 (0.30-0.52) | 0.59 (0.48-0.70) | 5.38 (4.99-5.75) | -0.36 (-0.45, -0.27) | 3.40 (2.76-4.36) | 0.60 (0.49-0.71) |
| **Late cART[24-59m]** | 0.54 (0.43-0.65) | 0.46 (0.35-0.57) | 5.68 (5.13-6.21) | -0.33 (-0.42, -0.23) | 4.18 (3.43-5.32) | 0.69 (0.53-0.81) |

cART: combination antiretroviral therapy; MCV: measles-containing vaccine. The analysis includes children with at least 2 observations and censors children at seroconversion (seronegative to positive) or at ≥ 4-fold increase in antibody levels. Posterior median and 95% credible intervals are presented**.**

[a] Early cART[0-8m] defined as cART initiation 0-8 month of age; Late cART[9-23m] defined as cART initiation 9-23 months of age; Late cART[24-59m] defined as cART initiation 24-59 months of age

**Supplemental Digital Content 11. Sensitivity analysis of the post-MCV1 antibody decay analysis including all children living with HIV with at least one observation to contribute to the analysis**

| cART group (k) | Probability of being a non-responder ($w_k$) | Probability of being a decayer ($1-w_k$) | ART group-specific intercept, among decayers ($\alpha_k$) | ART group-specific slope, among decayers ($\beta_k$) | Mean time to seroreversion, among decayers who are seropositive at the time of MCV1 receipt, years ($T_k$) | Proportion of decayers who are seropositive at the time of MCV1 receipt |
|---|---|---|---|---|---|---|
| **Early cART[0-8m]** | 0.51 (0.36-0.66) | 0.49 (0.34-0.64) | 6.13 (5.48-6.77) | -0.92 (-1.16, -0.67) | 1.71 (1.33-2.24) | 0.80 (0.63-0.91) |
| **Late cART[9-23m]** | 0.48 (0.39-0.57) | 0.52 (0.43-0.61) | 5.62 (5.27-5.98) | -0.52 (-0.60, -0.43) | 2.48 (2.11-2.98) | 0.68 (0.56-0.77) |
| **Late cART[24-59m]** | 0.55 (0.45-0.64) | 0.45 (0.36-0.55) | 5.93 (5.47-6.41) | -0.46 (-0.55, -0.38) | 3.12 (2.69-3.67) | 0.76 (0.63-0.86) |

cART: combination antiretroviral therapy; MCV: measles-containing vaccine. The analysis includes children with at least 1 observation and censors children at seroconversion (seronegative to positive) or at ≥ 2-fold increase in antibody response. Posterior median and 95% credible intervals are presented**.**

[a] Early cART[0-8m] defined as cART initiation 0-8 month of age; Late cART[9-23m] defined as cART initiation 9-23 months of age; Late cART[24-59m] defined as cART initiation 24-59 months of age

### Supplemental Digital Content 12. Correlates of seropositivity among children living with HIV

| | Total N | Seropositive N (%) | p-value[g] | Crude PR (95% CI) | Adjusted PR (95% CI) |
|---|---|---|---|---|---|
| Number of participants | 301 | 119 (39.5) | | | |
| **Characteristics at cART initiation** | | | | | |
| Age of cART initiation | | | | | |
| 0-8 months | 39 | 12 (30.8) | 0.20 | 0.83 (0.50, 1.40) | 1.03 (0.63, 1.67) |
| 9-23 months | 138 | 51 (37.0) | | REF | REF |
| 24-59 months | 124 | 33 (26.6) | | 0.72 (0.50, 1.04) | 0.92 (0.54, 1.57) |
| Sex | | | | | |
| Male | 154 | 48 (31.2) | 0.78 | REF | - |
| Female | 147 | 48 (32.7) | | 1.05 (0.75,1.46) | - |
| Orphan status[a] | | | | | |
| Not an orphan | 276 | 84 (30.4) | 0.07 | REF | REF |
| Single or double orphan | 25 | 12 (48.0) | | 1.58 (1.10,2.46) | 1.55 (1.07, 2.24) |
| SES percentile | | | | | |
| 0-25% | 184 | 59 (32.1) | 0.87 | REF | - |
| 26-50% | 95 | 29 (30.5) | | 0.95 (0.66, 1.38) | - |
| 51-100% | 22 | 8 (36.4) | | 1.13 (0.63, 2.05) | - |
| cART regimen backbone | | | | | |
| NVP | 143 | 46 (32.2) | 0.65 | REF | - |
| EFV | 103 | 30 (29.1) | | 0.91 (0.62, 1.33) | - |
| LPV/r | 55 | 20 (36.4) | | 1.13 (0.74, 1.73) | - |
| Severe immunosuppression[b] | | | | | |
| No | 111 | 36 (32.4) | 0.90 | REF | - |
| Yes | 145 | 46 (31.7) | | 0.98 (0.68, 1.40) | - |
| Height-for-age Z-score[c] | | | | | |
| None/mild stunting | 59 | 20 (33.9) | 0.19 | REF | - |
| Moderate stunting | 62 | 15 (24.2) | | 0.71 (0.40, 1.26) | - |
| Severe stunting | 111 | 42 (37.8) | | 1.12 (0.73, 1.72) | - |
| Weight-for-age Z-score[c] | | | | | |
| None/mild underweight | 154 | 50 (32.5) | 0.95 | REF | - |
| Moderate underweight | 65 | 21 (32.3) | | 1.00 (0.65, 1.51) | - |
| Severe underweight | 66 | 20 (30.3) | | 0.93 (0.61, 1.44) | - |
| **Characteristics at measles antibody testing** | | | | | |
| Adherence[d] | | | | | |
| ≤ 95% | 64 | 18 (28.1) | 0.40 | 0.83 (0.53, 1.29) | - |
| > 95% | 180 | 61 (33.9) | | REF | - |
| Severe immunosuppression[b] | | | | | |
| No | 251 | 78 (31.1) | 0.41 | REF | - |
| Yes | 20 | 8 (40.0) | | 1.29 (0.73, 2.27) | - |
| Viral suppression[e] | | | | | |
| No | 43 | 7 (16.3) | 0.03 | 0.50 (0.24, 1.01) | 0.60 (0.29, 1.22) |
| Yes | 177 | 58 (32.8) | | REF | REF |
| Height-for-age Z-score[c] | | | | | |
| None/mild stunting | 81 | 16 (19.8) | 0.03 | REF | REF |
| Moderate stunting | 82 | 27 (32.9) | | 1.67 (0.97, 2.85) | 1.49 (0.92, 2.40) |
| Severe stunting | 116 | 43 (37.1) | | 1.88 (1.14, 3.09) | 1.66 (1.05, 2.63) |
| Weight-for-age Z-score[c] | | | | | |
| None/mild underweight | 217 | 68 (31.3) | 0.95 | REF | - |
| Moderate underweight | 38 | 11 (29.0) | | 0.92 (0.54, 1.58) | - |
| Severe underweight | 28 | 9 (32.1) | | 1.03 (0.58, 1.82) | - |
| Calendar year[f] | | | | | |
| 2007-9 | 19 | 2 (10.5) | <0.0001 | REF | REF |
| 2010 | 53 | 7 (13.2) | | 1.25 (0.28, 5.53) | 1.44 (0.32, 6.48) |
| 2011 | 53 | 37 (69.8) | | 6.63 (1.77, 24.95) | 6.54 (1.66, 25.73) |
| 2012 | 53 | 9 (17.0) | | 1.61 (0.38, 6.83) | 1.58 (0.37, 6.73) |
| 2013 | 30 | 14 (46.7) | | 4.43 (1.13, 17.41) | 4.43 (1.15, 17.09) |
| 2014 | 21 | 4 (19.1) | | 1.81 (0.37, 8.81) | 2.09 (0.44, 9.99) |
| 2015 | 30 | 8 (26.7) | | 2.53 (0.60, 10.71) | 2.70 (0.63, 11.47) |

| | | | | | |
|---|---|---|---|---|---|
| 2016 | 13 | 1 (1.0) | | 0.73 (0.07, 7.28) | 0.80 (0.08, 7.88) |
| 2017 | 14 | 7 (50.0) | | 4.75 (1.15, 19.54) | 4.67 (1.16, 18.73) |
| 2018-20 | 15 | 7 (46.7) | | 4.43 (1.07, 18.35) | 4.54 (1.13, 18.17) |

cART: combination antiretroviral therapy; CI: confidence interval; EFV: efavirenz; LPV/r: lopinavir plus ritonavir; NVP: nevirapine; PR: prevalence ratio; REF: reference group; SES: socioeconomic status

[a] Single/double orphan defined as either or both the child's mother or father deceased, as determined by caregiver report.
[b] Based on WHO criteria for severe immunosuppression, using CD4 percentage and age at cART initiation.
[c] Based on WHO growth standards. None/mild defined as a z-score ≥-2; moderate defined as a z-score <-2 and ≥-3; severe defined
[d] Adherence was measured by pill count and categorized as ≤95% or >95% based on the minimum value of all drugs counted.
[e] Viral suppression defined as a viral load <400 copies/mL.
as a z-score <-3.
[f] 2007-2009 and 2018-2020 were combined due to small sample sizes (n<10) in those years.
[g] p-value based on Pearson's chi-square test.

**Supplemental Digital Content 13. Measles antibody seroprevalence by HIV status and calendar year among children included in the cross-sectional analysis[a]**

| | Children with HIV (n=301) | | Controls (n=911) | |
|---|---|---|---|---|
| | Total N | Seropositive N (%) | Total N | Seropositive N (%) |
| 2007-09 | 19 | 2 (10.5) | 14 | 3 (21.4) |
| 2010 | 53 | 7 (13.2) | 104 | 56 (53.9) |
| 2011 | 53 | 37 (69.8) | 132 | 76 (57.6) |
| 2012 | 53 | 9 (17.0) | 117 | 97 (82.9) |
| 2013 | 30 | 14 (46.7) | 94 | 82 (87.2) |
| 2014 | 21 | 4 (19.1) | 48 | 40 (83.3) |
| 2015 | 30 | 8 (26.7) | 192 | 164 (85.4) |
| 2016 | 13 | 1 (1.0) | 72 | 60 (83.3) |
| 2017 | 14 | 7 (50.0) | 101 | 92 (91.1) |
| 2018-20 | 15 | 7 (46.7) | 37 | 27 (73.0) |

[a] 2007-2009 and 2018-2020 were combined due to small sample sizes (n<10) in those years.

**Supplemental Digital Content 14. Complete individual-level trajectories from the 195 individuals who had at least one observed boost during follow-up**

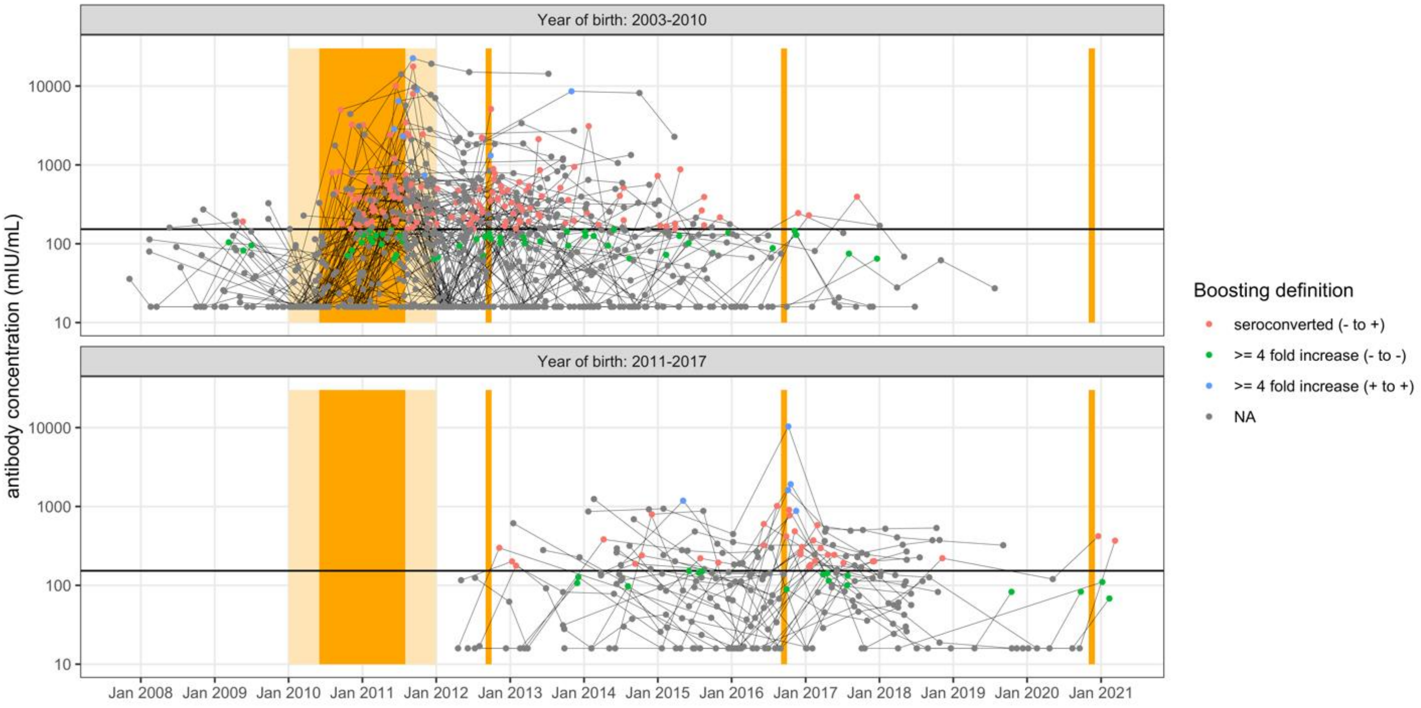


Boosts were defined as a seroconversion or a ≥4-fold increase in the antibody concentration between two time points. X-axis represents calendar date, and y-axis represents adjusted antibody concentration. The color of the circle represents the type of boosting observed at that time point (red: seroconverted from seronegative to seropositive, green: ≥4-fold increase from seronegative to seronegative, blue: ≥4-fold increase from seropositive to seropositive, grey: no boost). Each panel represents a group of birth cohorts (i.e., grouped by year of birth). The black horizontal line on each panel indicates the cutoff for seropositivity. The orange shading represents the timing of potential measles exposures: outbreak and SIA in June 2010-July 2011, SIA in September 2012, SIA in September 2016, and SIA in November 2020. The light orange shading represents the 5-month buffer before and after the outbreak period for which observations were included in the outbreak period boosting analysis.

**Supplemental Digital Content 15. Individuals with observed boosting episodes during follow-up and probability of observed boosting**

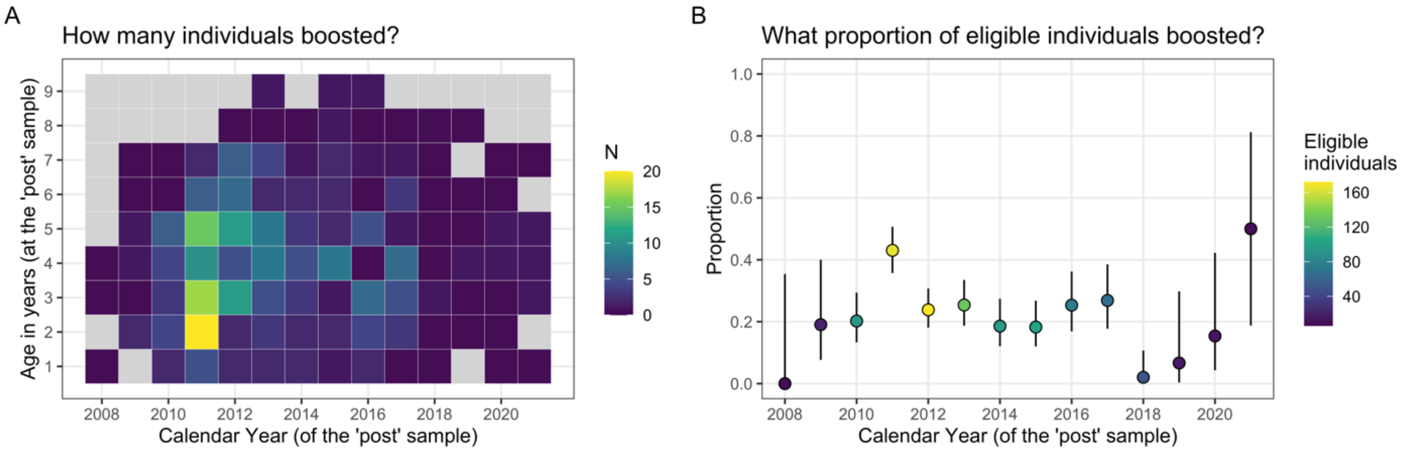


A boost is defined as a seroconversion or a ≥4-fold increase in the antibody titer between two time points (i.e., ‘pre’ and ‘post’ samples). **(A)** The number of individuals who experienced an antibody boost, stratified by the calendar year of the ‘post’ sample and by age in years at the ‘post’ sample. Gray squares indicate year-age strata with no individuals who could have had an antibody boost. **(B)** The proportion who experienced an antibody boost among all individuals who had a ‘post’ sample during a calendar year (i.e., ‘eligible individuals’) with 95% binomial confidence intervals.

**Supplemental Digital Content 16. Changes in measles antibody concentration during the 2010-2011 outbreak period**

| Pattern of antibody responses during outbreak period | Number of individuals | Percentage of individuals |
|---|---|---|
| No boosting | 82 | 49.1% |
| Boosted (-ve to -ve) | 12 | 7.2% |
| Boosted (+ve to +ve) | 4 | 2.4% |
| Seroconverted | 69 | 41.3% |

Patterns of antibody responses among the 167 individuals who had at least 2 observations between January 2010 and December 2011, and whose earliest time point was before August 2011 and latest time point was after June 2010. For the 82 individuals who were observed to have no boosting during this time, 54 were consistently seronegative, 20 were consistently seropositive, and 8 seroreverted.
